# Repurposing drugs for the prevention of vascular dementia: Evidence from drug target Mendelian randomization

**DOI:** 10.1101/2025.04.11.25325641

**Authors:** Victoria Taylor-Bateman, Phazha Bothongo, Venexia Walker, Patrick G Kehoe, Liv Tybjærg Nordestgaard, Yoav Ben-Shlomo, Neil M Davies, Dylan M Williams, Emma L Anderson

**Affiliations:** Division of Psychiatry, University College London, London, UK; Medical Research Council Integrative Epidemiology Unit, Bristol Medical School, University of Bristol, Bristol, Oakfield House, Oakfield Grove, BS8 2BN, United Kingdom (Liv); Department of Surgery, Perelman School of Medicine, University of Pennsylvania, Philadelphia, USA; Cerebrovascular and Dementia Research Group, Bristol Medical School, University of Bristol, Learning & Research, Southmead Hospital, Bristol, BS10 5NB; Department of Clinical Biochemistry, Copenhagen University Hospital - Herlev and Gentofte, Borgmester Ib Juuls Vej 1, 2730 Herlev, Denmark (Liv); Population Health Sciences, Bristol Medical School, University of Bristol, Bristol, 39 Whatley Road, BS8 2PZ, United Kingdom; Department of Statistical Science, University College London, London WC1E 6BT, UK; Department of Public Health and Nursing, Norwegian University of Science and Technology, Norway; Unit for lifelong health and ageing, University College London, London, UK

## Abstract

**Importance:** Vascular dementia (VaD) is a devastating cerebrovascular disease with no disease-modifying treatments available. Repurposing existing drugs for VaD risk factors could have an important clinical impact.

**Objective:** To determine whether lipid-lowering, anti-hypertensive, or anti-inflammatory drug targets affect the risk of vascular dementia using Mendelian randomization.

**Design:** Evidence suggests that higher cholesterol and blood pressure are associated with increased VaD risk, and inflammation is thought to play a key role in pathogenesis. Two-sample MR was conducted using *cis-*acting genetic variants in genes encoding each drug target, and data on five VaD-related outcomes. Instrument performance was assessed with positive controls (coronary artery disease, heart failure, stroke and rheumatoid arthritis).

**Setting:** Summary-level genetic data

**Participants:** Publicly available genetic association data from large cohorts of European ancestry. To maximize the sample size for vascular dementia risk as an outcome, we conducted a meta-analysis of case-control data from FinnGen & MEGAVCID.

**Exposures:** Genetically proxied drug effects for 46 lipid-lowering (n=17), antihypertensive (n=18), and anti-inflammatory (n=11) targets.

**Main Outcomes and Measures:** Odds ratios/betas and 95% CIs for VaD outcomes (clinical diagnosis, white matter hyperintensity volume, fractional anisotropy, mean diffusivity and lacunar stroke diagnosis) were estimated per 1-unit change in the exposure.

**Results:** For VaD risk, N=7,009 cases and N=899,672 controls were used. Neuroimaging outcome datasets included a maximum of N=50,559 participants. Beta-1 adrenergic receptor (ADRB1) was the only target for which there was consistent, albeit modest, evidence of benefit for four out of the five outcomes (clinical diagnosis: OR= 0.90, 95%CI 0.80 to 1.01, white matter hyperintensities: β= -0.03, 95%CI -0.07 to 0.00, mean diffusivity: β= -0.18, 95%CI -0.37 to 0.00, lacunar stroke: OR= 0.91, 95%CI 0.80 to 1.03). Angiotensin-converting enzyme (ACE) inhibition was suggested to increased VaD risk (OR= 1.12, 95%CI 1.01 to 1.24). There was little evidence to suggest other targets affect the outcomes.

**Conclusions and relevance:** ARDB1 antagonism may be a promising repurposing candidate for VaD. Pharmacovigilance studies are required to further examine ACE inhibitors’ potential to increase VaD risk. There is little evidence to support repurposing of many licensed lipid-lowering, antihypertensive and anti-inflammatory drugs for VaD prevention or treatment.

**KEY POINTS:** *Question:* Can existing drugs with potential neurovascular benefits (lipid-lowering, antihypertensive, and anti-inflammatory therapies) be repurposed for vascular dementia (VaD) treatment/prevention?

*Findings:* We used large genetic datasets for VaD and its neuroimaging features in drug target Mendelian randomization. ARDB1 antagonist exposure was associated with lower VaD risk and better neuroimaging phenotypes. Conversely, exposure to ACE inhibitors may increase VaD risk. There was little evidence for other drug targets.

*Meaning:* ADRB1 antagonists could be promising candidates for drug repurposing for VaD prevention and treatment. Pharmacovigilance research is warranted to confirm or refute a link between ACE inhibitor use and VaD risk.

## INTRODUCTION

More than 55 million people are living with dementia globally, but very few effective treatments are available. Evidence of cerebrovascular disease is present in almost 80% of all dementia cases^1^; and there is some evidence that it might be one of the earliest pathological changes that occurs in Alzheimer’s disease^2^, which is still widely reported as the leading cause of dementia. Yet, vascular dementia (VaD), has received comparatively little attention and research funding^3^. VaD is a progressive cerebrovascular disease caused by brain injury as a result of impaired cerebral blood flow^4^. There are currently no effective treatments or preventive therapeutics for VaD; options are limited to modifying risk factors such as cholesterol levels and blood pressure and managing symptoms.

Drug repurposing can reduce both the time and cost associated with introducing new treatments, while leveraging established safety profiles and existing regulatory approvals. It has been widely explored in the case of Alzheimer’s disease with over 573 drugs proposed as therapeutic candidates in the last 10 years^5^. However, research into drug repurposing for VaD is scarce^6, 7, 8, 9^ and current findings are limited. In addition to most drug trials being very small (n<500), they have also been restricted to participants of old age (e.g. >70 years). This is problematic since the onset of prodromal cerebrovascular pathology begins in midlife^10^, and any preventative interventions are likely to be more effective when administered as early as possible in the disease course. Yet, conducting clinical trials in which preventative drugs are initiated in midlife, with sufficiently long clinical follow-up periods to detect VaD outcomes, is not feasible nor cost-effective.

Genetic epidemiology provides another source of evidence about the potential effects of licenced and new drugs, and evidence has shown that therapeutics with human genetic support are more than twice as likely to achieve regulatory approval than agents lacking genetic support^11^. Drug target Mendelian randomization (MR) is an instrumental variable method that can be used to identify potential high-priority therapeutic targets^12^. It has already been successfully applied in other disease areas: for example, to support the repurposing of interleukin-6 receptor inhibitors to treat severe COVID-19 infection^13, 14^. In drug target MR, the expression or function of protein drug targets are instrumented by genetic variants within or near the genes that encode them. These genetic effects are then used to estimate the effects of the drug target on a disease or proxy endpoint. Genotypes are inherited randomly during conception, analogous to random treatment allocation in randomized trials. Thus, associations of genetic markers for both drug targets and disease outcomes are unlikely to be biased by confounding or reverse causation, which limits causal inference in traditional observational epidemiology methods. With the wealth of publicly available genetic data, this method represents an efficient and cost-effective approach to identifying novel druggable targets or evaluating the repurposing potential of existing drugs.

Prior studies have suggested that blood pressure and low-density lipoprotein cholesterol (LDL-c) are causal risk factors for VaD^15^.Systemic inflammation is also thought to be an important part of the disease’s pathogenesis with higher C-reactive protein (CRP) levels having been previously associated with VaD neuroimaging markers^16^. This study aimed to evaluate the potential for repurposing a range of anti-hypertensive, lipid-lowering, and anti-inflammatory drugs to reduce risk of VaD using two-sample drug target MR.

## METHODS

### Drug target selection

Figure 1 provides an overview of the data and methods used in our study. For each class of medication considered (lipid-lowering, antihypertensive, and anti-inflammatory), the key drug targets of licensed therapeutics were identified using a combination of the BNF (https://bnf.nice.org.uk/) and NICE (www.nice.org.uk) guidelines. Our search resulted 46 potential targets (Table 1). We identified each drug’s target using https://go.drugbank.com/^17^ and the coordinates of protein-coding gene region(s) determined using https://www.ncbi.nlm.nih.gov/gene^18^.

**Table 1:**
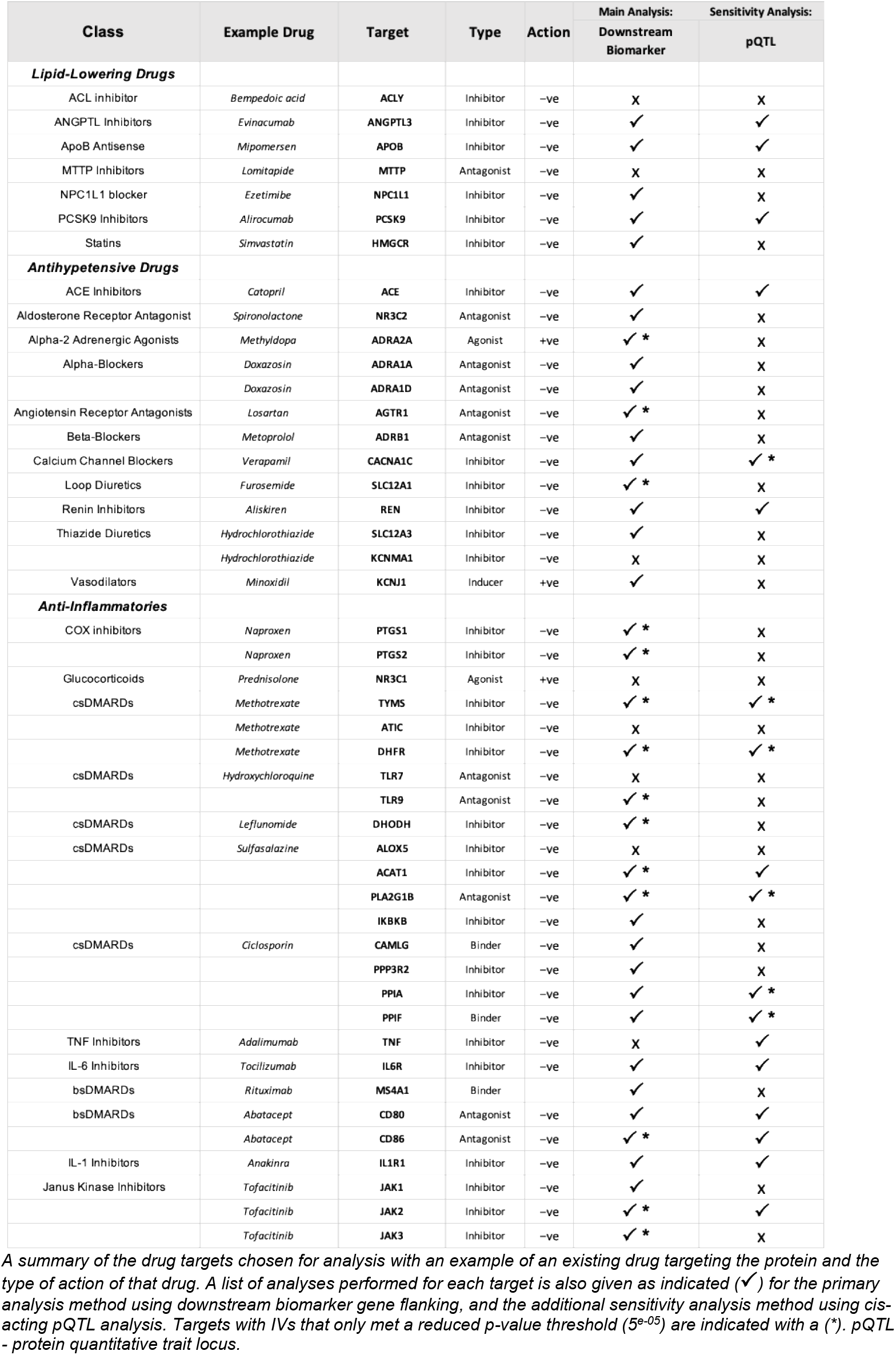
A list of the identified drug targets by class and which analysis was performed.

**Figure 1:**
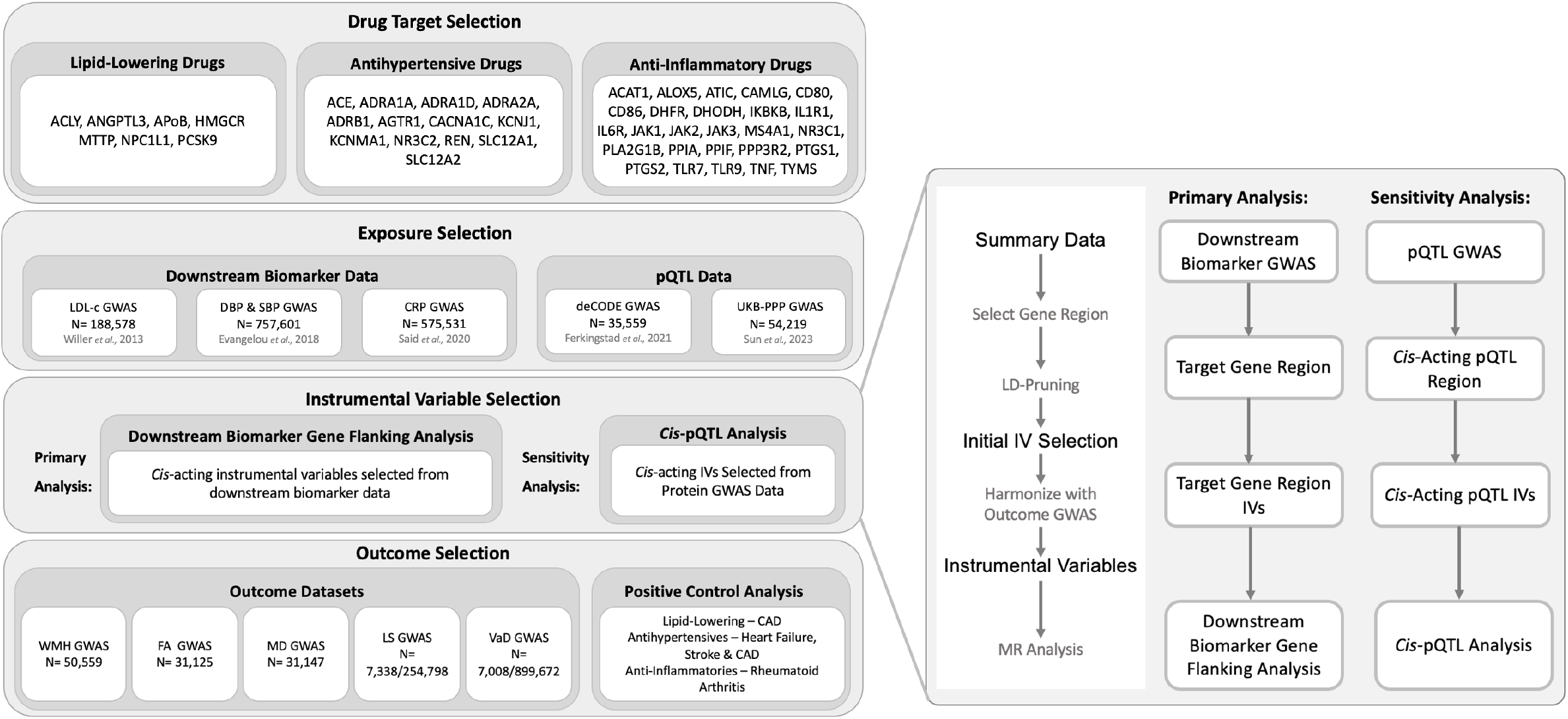
Illustration of data, instrumental variable selection and analyses performed. A flowchart summarizing the methods used in this analysis including drug-target selection, exposure and outcome selection and obtaining instrumental variables (IVs). GWAS (genome-wide association study), pQTL (protein quantitative trait locus), LDL-C (Low-density lipoprotein cholesterol), DBP (Diastolic blood pressure), SBP (Systolic blood pressure), CRP (C-reactive protein), WMH (White matter hyperintensities), FA (Fractional anisotropy), MD (Mean Diffusivity), LS (Lacunar stroke), VaD (Vascular Dementia), CAD (coronary artery disease).

### Data

#### Outcome Data

We considered five different VaD outcomes: VaD clinical diagnosis, white matter hyperintensity volume (WMH), fractional anisotropy (FA), mean diffusivity (MD) and lacunar stroke. For VaD clinical diagnosis, we conducted a meta-analysis of two existing case-control GWAS using METAL (full details in the online supplement)^19^, combining summary data from MEGAVCID *et al*., 2024 (N cases= 3,892, N controls= 466,606)^20^ and the FinnGen study^21^ (N cases= 3,116, N controls= 433,066). This gave a maximum of N= 7,009 cases and N= 899,672 controls. We also considered four neuroimaging features of VaD which are used in clinic for diagnosis and prognosis: WMH, MD, FA, and lacunar stroke. For ease of interpretation, the directionality of FA was inversed (called iFA throughout) to match the direction of the other VaD outcomes. Thus, higher values of all traits represent worse cerebrovascular health. Summary-level GWAS data for these traits were obtained from the largest, most recent studies: WMH (N= 50,559)^22^; FA and MD (N=31,125 and 31,147, respectively)^15^; and lacunar stroke (N= 7338 cases, 254,798 controls)^23^. A smaller WMH GWAS dataset (N= 31,855)^15^ was used if genome-wide significant variants for drug targets were not available in the primary WMH meta-analysis.

#### Downstream Biomarker Data

GWAS summary data information can be found in Supplementary Table 1. GWAS summary data were obtained for four downstream biomarkers (i.e. traits expected to be targeted by each drug). These included; LDL-C for lipid-lowering targets (Willer *et al*., 2013, n=188,578)^24^; systolic and diastolic blood pressure (SBP and DBP) (Evangelou *et al*., 2018, n=757,601) for antihypertensive targets^25^, and CRP for anti-inflammatory targets (Said *et al*., 2022, n=575,531)^26^. The interpretation of results is per unit increase in the downstream biomarker (i.e., per 1 mmHg increase for SBP and DBP, per SD increase for LDL-C, and per 1 unit increase in natural log-transformed CRP). A list of chosen instrumental variables can be found in Supplementary Table 2.

#### Positive control data

We evaluated instrument validity using positive control traits (i.e., disorders for which each drug is licenced to treat) as additional outcomes. For lipid-lowering drug targets, we considered coronary artery disease (CAD) (N cases= 122,733, N controls= 424,528)^27^; for antihypertensive targets we considered heart failure (N cases= 22,350, N controls= 74,823)^28^, stroke (N cases= 67,162, N controls= 454,450)^29^ and CAD^27^; for anti-inflammatory targets we considered rheumatoid arthritis (RA) (N cases= 22,350, N controls= 74,823)^30^.

#### Statistical Analyses

All analyses were conducted using R (version 4.4.0). For each drug target, *cis*-acting genetic variants (within a 500kb region on either side of the target gene), which were associated with the downstream biomarker at genome-wide significance (p< 5×10^−8^), were identified^31^.

Independent instruments were selected using an LD clumping threshold of r^2^ < 0.001 within a 10,000kb distance. The P-value threshold was lowered if no instruments were identified at genome-wide significance (p< 5×10^−5^). It is worth noting that there are limitations to lowering the P-value threshold, as it increases the risk of using potentially invalid instruments and horizontal pleiotropy, which may bias our results.

All effect alleles were aligned to harmonize data across the drug target and VaD outcome datasets^31^. The number of instrumental variables used for each analysis and the corresponding F-statistics, which measure instrument strength, were calculated. Targets with an F statistic below 10 were interpreted to be susceptible to weak instrument bias. Two-sample MR was performed using a Wald-ratio approach (if only one genetic instrument was available) or an inverse-variance weighted (IVW) (if more than one genetic instrument was available) as the primary analysis with the ‘MendelianRandomization; R package^32, 33^. Full details of the Wald-ratio and IVW methods are in the supplement. Where the number of instrumental variables allowed, Cochran’s Q statistics and additional MR sensitivity analyses were calculated to examine potential bias due to horizontal pleiotropy (weighted median MR^34^ & MR-Egger^35^, full details in supplement). At least one positive control was evaluated as an outcome for each drug class (coronary artery disease for lipid-lowering drugs; heart failure, CAD, and stroke for antihypertensive drugs; and rheumatoid arthritis for anti-inflammatory drugs) to validate instrument validity.

#### Sensitivity analyses

Several sensitivity analyses were conducted including: replicating results using protein Quantitative Trait Loci (pQTLs), instead of downstream biomarkers, as the exposure genetic associations; a trans-acting MR analysis; MRlap to account for sample overlap; statistical colocalization; alternative downstream biomarker selection; and alternative positive controls for anti-inflammatory targets. Full details of all sensitivity analyses are described in the supplement.

## RESULTS

Of the 46 drug targets identified, suitable instruments were identified for 38 targets in the downstream biomarker GWAS data (all except for ACLY, ALOX5, ATIC, KCNMA1, MTTP, NR3C1, TLR7 and TNF, Table 1). Where an analysis was not performed, this was due to either data not being present or a lack of suitable instruments meeting either genome-wide significance or a reduced p-value threshold of p< 5×10^−5^.

### Lipid-lowering targets

Figure 2 shows the results for lipid-lowering targets for which there was evidence of an effect in the expected direction on the positive control, CAD. Full results for all targets are in Supplemental Table 3 and Supplemental Figure 1. Overall, there was little consistent evidence suggesting lipid-lowering drug targets had a causal effect on VaD. There was evidence that HMGCR inhibition may reduce the risk of lacunar stroke (OR= 0.50, CI: 0.29 to 0.87), and weak evidence that it may reduce VaD risk. However, the VaD estimate lacked precision, despite the point estimate being larger than that for HMGCR on CAD. NPC1L1 increased iFA (β= 1.65, CI: 0.02 to 3.28). There was little evidence for the effect of these targets on any other outcomes.

**Figure 2:**
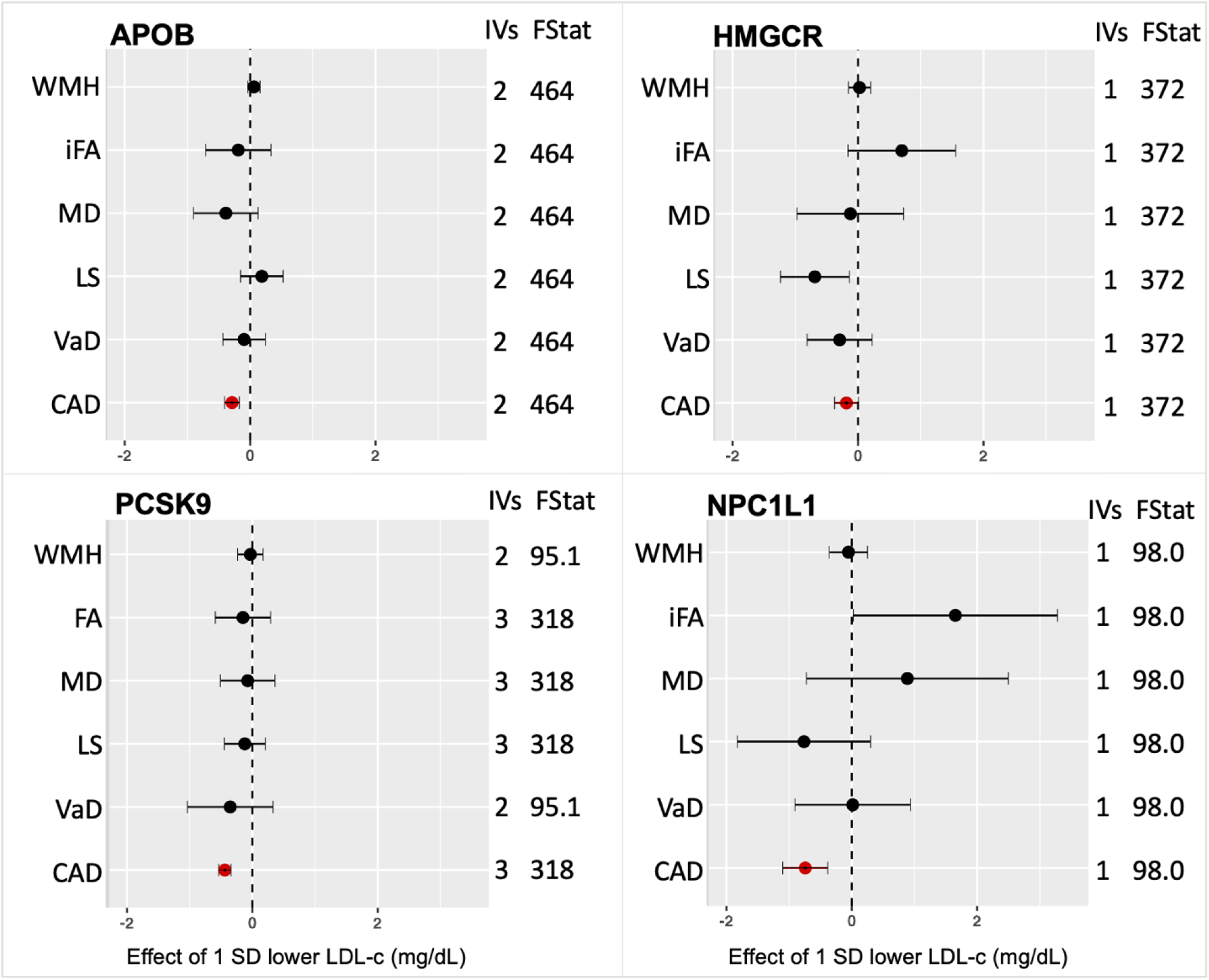
Drug target MR for lipid-lowering drug targets. MR was conducted for lipid-lowering drug targets using LDL-C (a downstream biomarker) as the exposure with five vascular dementia outcomes; WMH (white matter hyperintensities), iFA (inversed fractional anisotropy), MD (mean diffusivity), LS (lacunar stroke), VaD (vascular dementia diagnosis). CAD (coronary artery disease) was used as a positive control and is indicated in red. The number of instrumental variables (IVs) used in each analysis is given along with the F-statistic (Fstat). LDL-C (low-density lipoprotein cholesterol), SD (standard deviation). Targets lacking evidence of a reductive effect on the positive control are presented in the online supplement (Supplementary figure 1.).

### Anti-hypertensive targets

Figure 3 shows the results of antihypertensive targets for which there was evidence of an effect in the expected direction for at least one of the positive controls, stroke, heart failure, and CAD. Results for all other targets with little evidence of an effect on the positive controls are presented in Supplemental Figures 2 (weighted by DBP) and 3 (weighted by SBP). Results were broadly consistent when using DBP or SBP as the downstream biomarker. There was consistent evidence to support a protective effect of ADRB1 antagonists on lower WMH (β= -0.03, CI: - 0.07 to 0.001), lower MD (β= -0.18, CI: -0.37 to 0.004), lower VaD risk (OR= 0.90, CI: 0.80 to 1.01) and lower lacunar stroke risk (OR=0.91, CI: 0.80 to 1.03). There was also evidence of a protective effect of renin on iFA (β= -0.53, CI: -0.89 to -0.18) and MD (β= -0.42, CI: -0.77 to - 0.07) but no other outcomes. For those targets that could only be run using SBP, lacunar stroke risk was lower with both AGTR1 (OR= 0.77, CI: 0.59 to 1.00) and ADRA2A (OR= 0.66, CI: 0.48 to 0.90) modulation, but these targets were not indicated to affect other outcomes. Unexpectedly, there was evidence to suggest ACE inhibition increased risk of VaD; point estimates for WMH, MD and FA were also in the risk-increasing direction but lacked precision. The VaD risk-increasing estimates for ACE were highly consistent and were replicated in both the SBP and DBP downstream biomarker MR, as well as both pQTL sensitivity analysis MRs (in both deCODE and UKB-PPP).

**Figure 3:**
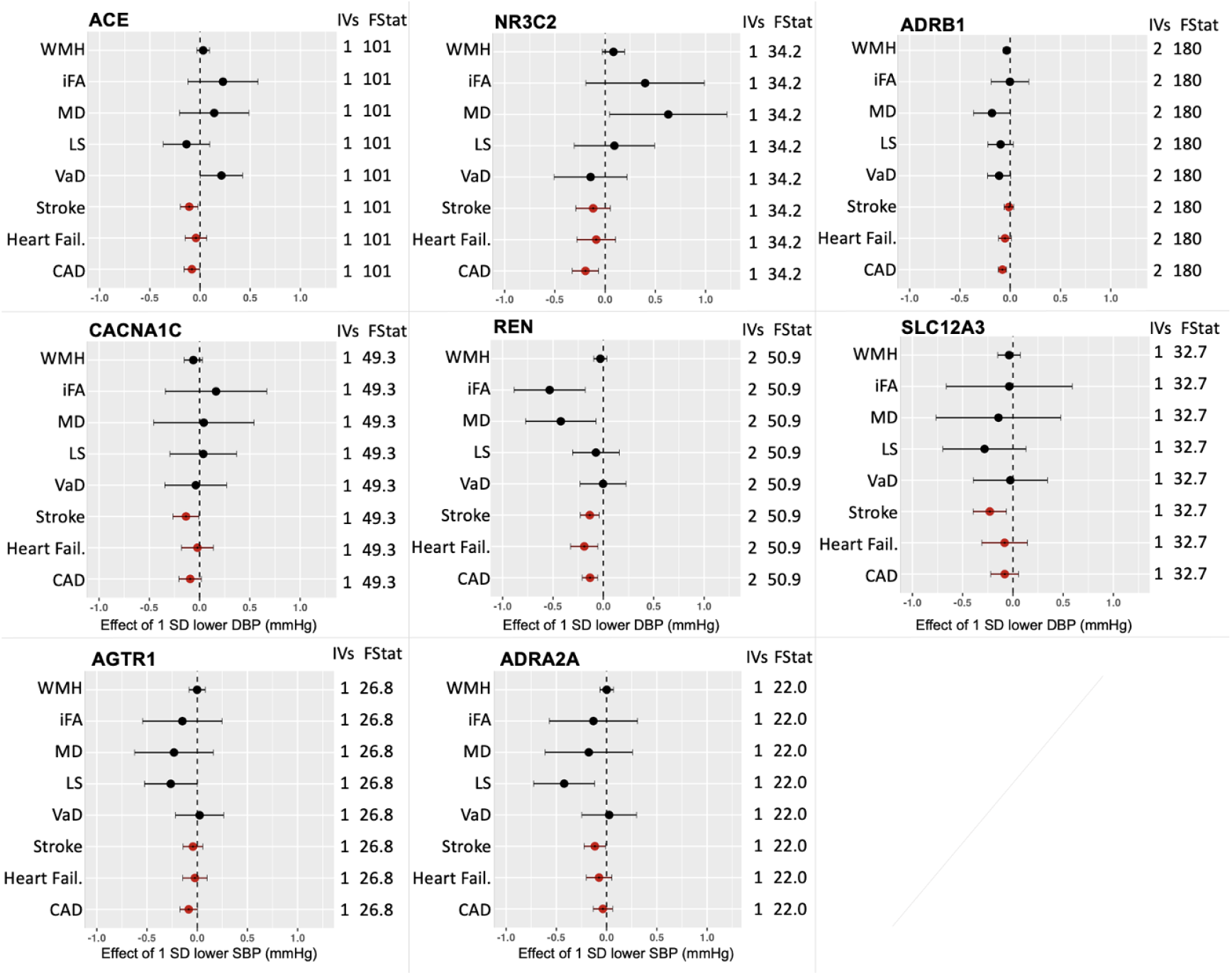
Drug target MR for anti-hypertensive drug targets. MR was conducted for anti-hypertensive drug targets using blood pressure (a downstream biomarker) as the exposure with five vascular dementia outcomes; WMH (white matter hyperintensities), iFA (inversed fractional anisotropy), MD (mean diffusivity), LS (lacunar stroke), VaD (vascular dementia diagnosis). This was conducted with both DBP (diastolic blood pressure) and SBP (systolic blood pressure), only DBP results have been present except for cases when no DBP analysis could be conducted (ADRA2A and AGTR1). CAD (coronary artery disease), stroke, and heart failure (Heart Fail.) were used as a positive control and are indicated in red. Targets where the positive control is in the unexpected direction or with no control with a p-value < 0.1 have been excluded. The number of instrumental variables (IVs) used in each analysis is given along with the F-statistic (Fstat). SD (standard deviation)

### Anti-inflammatory targets

Figure 4 shows the results of anti-inflammatory targets for which there was evidence of an effect in the expected direction for the positive control outcome, RA. Results for all other targets with little evidence of an effect on this primary positive control outcome are presented in Supplemental Figures 4a-c. Out of the 21 anti-inflammatory targets with suitable instruments identified in the downstream biomarker (CRP) GWAS, only three targets showed evidence of an expected effect on RA despite all targets being approved for the treatment of RA. Overall, there was little consistent evidence to suggest a causal effect of any of the anti-inflammatory drug targets on VaD endpoints. There was suggestive evidence that both IL1R1 and IL6R reduced lacunar stroke risk, but confidence intervals crossed the null. JAK2 also increased the risk of VaD, but there was no evidence of an effect on any other outcomes.

**Figure 4:**
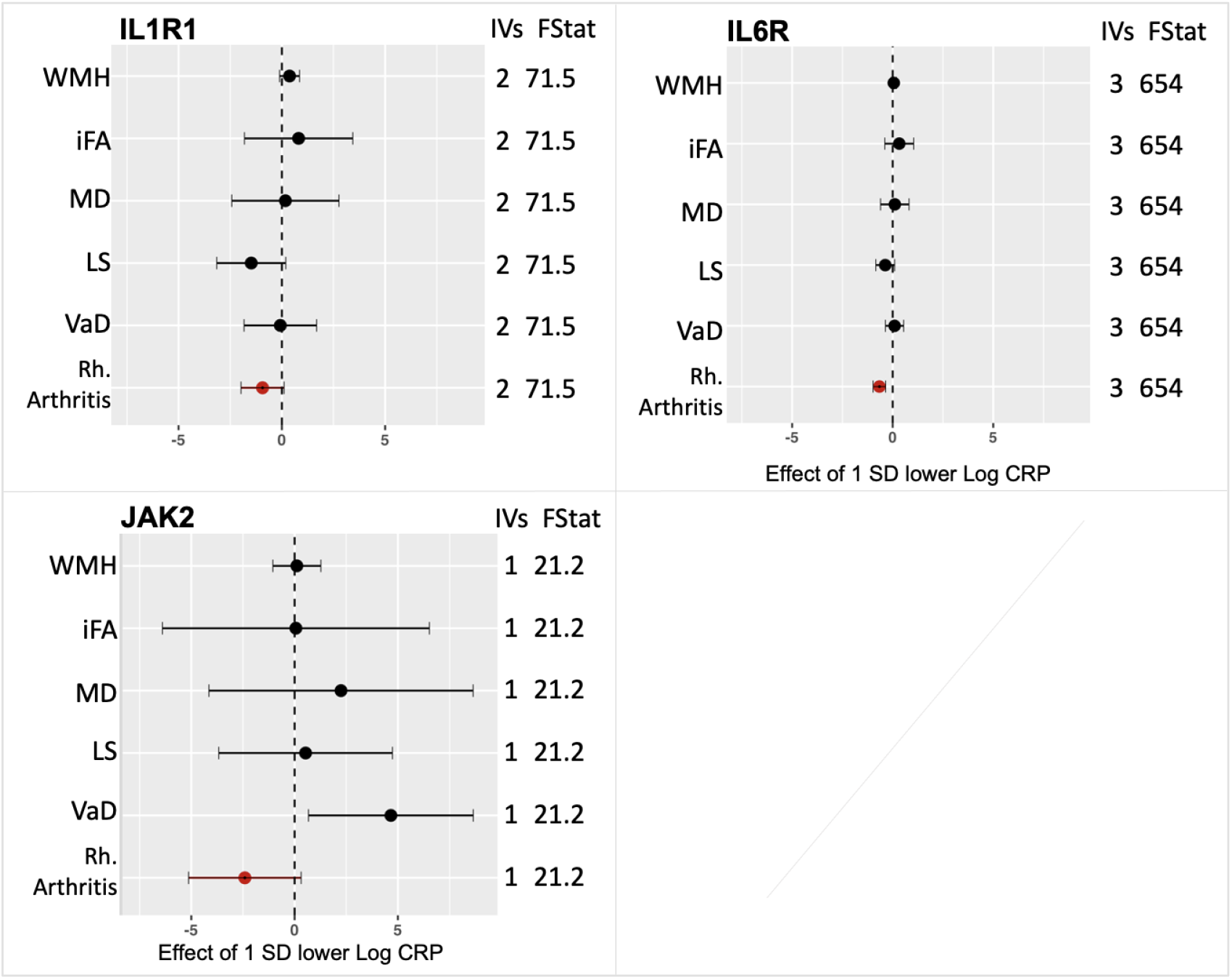
Drug target MR for anti-inflammatory drug targets. MR was conducted for anti-inflammatory drug targets using a downstream biomarker (CRP (C-reactive protein)) and five vascular dementia outcomes; WMH (white matter hyperintensities), iFA (inversed fractional anisotropy), MD (mean diffusivity), LS (lacunar stroke) and VaD (vascular dementia diagnosis). Rheumatoid arthritis (Rh. arthritis) was used as a positive control and is indicated in red. The results for all targets with a positive control in the expected direction and with a p-value < 0.1 are presented. The number of instrumental variables (IVs) used in each analysis is given along with the F-statistic (Fstat). SD (standard deviation).

### Sensitivity analyses

Complete results of all sensitivity analyses are in the supplement (Tables 3 – 10 and Figures 1 – 10). Overall, results were broadly comparable in all sensitivity analyses, and none significantly changed the conclusions of our study.

## DISCUSSION

Our results suggest limited opportunities for repurposing many existing lipid-lowering, antihypertensive, and anti-inflammatory drugs for treating VaD. There was, however, consistent evidence for ADRB1 antagonists, which were suggested to affect VaD risk, lacunar stroke risk, WMH volume, and MD. Genetically indexed ACE inhibition was, unexpectedly, associated with worse cerebrovascular outcomes. There are also some drug classes, such as HMGCR inhibition, for which our results were ambiguous and clearly warrant further follow-up in relation to cerebrovascular diseases.

ADRB1 is a G protein-coupled receptor that binds to epinephrine and norepinephrine, leading to physiological effects such as increased heart rate, contractility, and cardiac output^36^. ADRB1 is expressed in several brain regions, including the prefrontal cortex and the hippocampus, which play an important role in memory and learning^37^. Although there is a well-established relationship between hypertension and VaD^15^ there remains limited evidence specifically linking beta-blockers to neurological diseases. One study found that patients with hypertension taking beta-blockers capable of crossing the blood-brain barrier had a lower incidence of Alzheimer’s disease compared with those on less permeable beta-blockers^38^, and another found that patients with hypertension who were treated with beta-blockers had better cognitive performance compared with those treated with other antihypertensive medications^39^. An instrumental variable analysis using prescribers’ preference as the instrument also reported beta-blockers conferred small protective effects on dementia risk (13 fewer cases [95% CI= 6, 20] of any dementia per 1000 treated, compared with other antihypertensives). However, none of these studies examined the effects on VaD specifically. Given the lack of evidence for other blood-pressure modulating targets, it is plausible that any protective effects of ARDB1 antagonists on VaD risk act through mechanisms independent of blood pressure modulation.

Our finding that ACE inhibition was linked to higher VAD risk seems counterintuitive, particularly given (i) we observed a protective effect on all-stroke risk, and (ii) the strong association of blood pressure with increased VaD risk, and the effectiveness of ACE inhibitors for reducing blood pressure. ACE inhibitors lower blood pressure by preventing the production of angiotensin II, a hormone that narrows blood vessels and increases blood volume^40^. Similar adverse effects of ACE inhibitors have been observed previously for dementia risk, both in an observational study with all-cause dementia as an outcome^41^, and in two MR studies with Alzheimer’s disease as an outcome^42, 43^. Combined, these findings suggest that ACE inhibition may potentially increase dementia risk – possibly via mechanisms not involving blood pressure modulation, given that few effects were observed for other blood pressure modulating targets. It is also worth noting that AGTR1, another target which also inhibits angiotensin II (by binding to and blocking angiotensin II receptors, rather than inhibiting angiotensin II production like ACE), did not show evidence of an effect on VaD risk.

### Strengths and limitations

Our study provides the most comprehensive analysis of lipid-lowering, antihypertensive and anti-inflammatory drug targets for preventing or treating VaD. We used the largest GWAS available and conducted a replication pQTL analysis, triangulating findings across clinical diagnoses and neuroimaging endophenotypes. Our drug target MR study design enabled us to avoid the most ubiquitous limitations of previous observational studies of medication use: confounding by indication and reverse causation. This increases confidence that the effects we observe are causal, and may refute observational associations for targets where genetic evidence is null.

There are several limitations to our study. Firstly, choosing a suitable downstream biomarker for certain drug targets can be challenging. For example, it is unclear that CRP is the most appropriate biomarker for anti-inflammatory drugs (we chose CRP due to its use as a marker of systemic inflammation^26^). In the alternative biomarker sensitivity analyses, we considered IL-6; however, no genetic instruments were identified for any target. Secondly, the performance of positive controls was inconsistent across antihypertensive and anti-inflammatory targets, and many targets were discounted due to positive controls not performing well. It is challenging to unpick whether this is because the genetic variants were not relevant and there was a lack of statistical power (a violation of MR assumption of IV1: relevance), or that we did not select the most appropriate positive control for each target. Anti-inflammatory drugs are used to treat many conditions, so a tailored approach to each target is required. RA was the only disease for which all targets were approved to treat in our study; only a small number of targets were approved for treating Crohn’s disease and ulcerative colitis. In other cases, the relationship between specific drug targets and the positive controls may be complex. For example, loop diuretics are prescribed to treat complex hypertension. Still, previous research has shown that in some cases, they may actually worsen CAD risk^44^. Third, for some targets, there is sample overlap for some analyses. However, the MRlap sensitivity analysis correcting for bias due to sample overlap was broadly consistent. Fourth, we did not look at any drug-drug interactions, and it is plausible that effects on VaD outcomes may differ if a combination of drug classes are simultaneously targeted (e.g. by taking an antihypertensive and a statin). Fifth, our estimates are only relevant to specific target modulation and do not incorporate off-target effects of specific therapeutic agents. Sixth, whilst the largest datasets available were chosen, sample sizes were still relatively small, particularly for clinically diagnosed VaD, potentially limiting statistical power. Seventh, all neuroimaging biomarker GWASs primarily consisted of UK Biobank participants, thus, average ages of the samples were relatively young for dementia pathology. We may have observed different findings using a GWAS of an older population, but as yet, these do not exist. Lastly, all analyses were restricted to European ancestry samples due to data availability, and results may not generalise to other ancestry groups.

## CONCLUSIONS

ARDB1 antagonists appear to be a promising candidate for potential repurposing; future studies should examine the mechanisms through which ARDB1 modulates VaD risk. ACE inhibitors may increase vascular (and other) dementia risk and pharmacovigilance studies are required to confirm this effect. There was little evidence that other lipid-lowering, antihypertensive, or anti-inflammatory drugs could be repurposed to prevent or treat VaD.

## Data Availability

All data produced in the present work are contained in the manuscript

https://doi.org/10.5281/zenodo.15190763.

## Data and code availability

Summary statistics were obtained from the original papers as listed in Supplementary Table 1. Code is available at https://doi.org/10.5281/zenodo.15190763.

## Contribution statement

ELA conceived the study, supervised all statistical analyses and contributed to writing all drafts of the paper. VTB conducted all statistical analyses and contributed to writing all drafts of the paper. DMW, NMD, VMW, LTN, YBS, PK, SL, MT and PB provided critical comments on and edits to the paper.

## References

1. Neuropathology Group. Medical Research Council Cognitive Function and Aging Study. Pathological correlates of late-onset dementia in a multicentre, community-based population in England and Wales. Neuropathology Group of the Medical Research Council Cognitive Function and Ageing Study (MRC CFAS). Lancet. 2001;357(9251):169–175. doi:10.1016/s0140-6736(00)03589-3

2. Iturria-Medina Y, Sotero RC, Toussaint PJ, Mateos-Pérez JM, Evans AC. Early role of vascular dysregulation on late-onset Alzheimer’s disease based on multifactorial data-driven analysis. Nat Commun. 2016;7:11934. doi:10.1038/ncomms11934

3. British Heart Foundation and UK Dementia Research Institute announce Centre for Vascular Dementia Research. Accessed March 21, 2025. https://www.bhf.org.uk/what-we-do/news-from-the-bhf/news-archive/2023/november/bhf-and-uk-dri-announce-centre-for-vascular-dementia-research

4. Sinha K, Sun C, Kamari R, Bettermann K. Current status and future prospects of pathophysiology-based neuroprotective drugs for the treatment of vascular dementia. Drug Discovery Today. 2020;25(4):793–799. doi:10.1016/j.drudis.2020.01.003

5. Grabowska ME, Huang A, Wen Z, Li B, Wei WQ. Drug repurposing for Alzheimer’s disease from 2012–2022—a 10-year literature review. Front Pharmacol. 2023;14. doi:10.3389/fphar.2023.1257700

6. en Dam VH, van den Heuvel DMJ, van Buchem MA, et al. Effect of pravastatin on cerebral infarcts and white matter lesions. Neurology. 2005;64(10):1807–1809. doi:10.1212/01.WNL.0000161844.00797.73

7. Yamano S, Horii M, Takami T, et al. Comparison between angiotensin-converting enzyme inhibitors and angiotensin receptor blockers on the risk of stroke recurrence and longitudinal progression of white matter lesions and silent brain infarcts on MRI (CEREBRAL study): rationale, design, and methodology. Int J Stroke. 2015;10(3):452–456. doi:10.1111/ijs.12085

8. Kopczak A, Stringer MS, Brink H van den, et al. Effect of blood pressure-lowering agents on microvascular function in people with small vessel diseases (TREAT-SVDs): a multicentre, open-label, randomised, crossover trial. The Lancet Neurology. 2023;22(11):991–1004. doi:10.1016/S1474-4422(23)00293-4

9. Wardlaw JM, Debette S, Jokinen H, et al. ESO Guideline on covert cerebral small vessel disease. Eur Stroke J. 2021;6(2):CXI–CLXII. doi:10.1177/23969873211012132

10. d’Arbeloff T, Elliott ML, Knodt AR, et al. White matter hyperintensities are common in midlife and already associated with cognitive decline. Brain Commun. 2019;1(1):fcz041. doi:10.1093/braincomms/fcz041

11. King EA, Davis JW, Degner JF. Are drug targets with genetic support twice as likely to be approved? Revised estimates of the impact of genetic support for drug mechanisms on the probability of drug approval. PLoS Genet. 2019;15(12):e1008489. doi:10.1371/journal.pgen.1008489

12. Gill D, Georgakis MK, Walker VM, et al. MR for studying the effects of perturbing drug targets. Wellcome Open Res. 2021;6:16. doi:10.12688/wellcomeopenres.16544.2

13. The REMAP-CAP Investigators. Interleukin-6 Receptor Antagonists in Critically Ill Patients with Covid-19. New England Journal of Medicine. 2021;384(16):1491–1502. doi:10.1056/NEJMoa2100433

14. Bovijn J, Lindgren CM, Holmes MV. Genetic variants mimicking therapeutic inhibition of IL-6 receptor signaling and risk of COVID-19. The Lancet Rheumatology. 2020;2(11):e658–e659. doi:10.1016/S2665-9913(20)30345-3

15. Taylor-Bateman V, Gill D, Georgakis MK, et al. Cardiovascular Risk Factors and MRI Markers of Cerebral Small Vessel Disease: A MR Study. Neurology. 2022;98(4):e343–e351. doi:10.1212/WNL.0000000000013120

16. van Dijk EJ, Prins ND, Vermeer SE, et al. C-Reactive Protein and Cerebral Small-Vessel Disease: The Rotterdam Scan Study. Circulation. 2005;112(6):900–905. doi:10.1161/CIRCULATIONAHA.104.506337

17. Knox C, Wilson M, Klinger CM, et al. DrugBank 6.0: the DrugBank Knowledgebase for 2024. Nucleic Acids Research. 2024;52(D1):D1265–D1275. doi:10.1093/nar/gkad976

18. Brown GR, Hem V, Katz KS, et al. Gene: a gene-centered information resource at NCBI. Nucleic Acids Res. 2015;43(Database issue):D36–42. doi:10.1093/nar/gku1055

19. Willer CJ, Li Y, Abecasis GR. METAL: fast and efficient meta-analysis of genomewide association scans. Bioinformatics. 2010;26(17):2190–2191. doi:10.1093/bioinformatics/btq340

20. The Mega Vascular Cognitive Impairment and Dementia (MEGAVCID) consortium. A genome-wide association meta-analysis of all-cause and vascular dementia. Alzheimer’s & Dementia. 2024;20(9):5973–5995. doi:10.1002/alz.14115

21. Kurki MI, Karjalainen J, Palta P, et al. FinnGen provides genetic insights from a well-phenotyped isolated population. Nature. 2023;613(7944):508–518. doi:10.1038/s41586-022-05473-8

22. Sargurupremraj M, Suzuki H, Jian X, et al. Cerebral small vessel disease genomics and its implications across the lifespan. Nat Commun. 2020;11(1):6285. doi:10.1038/s41467-020-19111-2

23. Traylor M, Persyn E, Tomppo L, et al. Genetic basis of lacunar stroke: a pooled analysis of individual patient data and genome-wide association studies. Lancet Neurol. 2021;20(5):351–361. doi:10.1016/S1474-4422(21)00031-4

24. Willer CJ, Schmidt EM, Sengupta S, et al. Discovery and refinement of loci associated with lipid levels. Nat Genet. 2013;45(11):1274–1283. doi:10.1038/ng.2797

25. Evangelou E, Warren HR, Mosen-Ansorena D, et al. Genetic analysis of over 1 million people identifies 535 new loci associated with blood pressure traits. Nat Genet. 2018;50(10):1412–1425. doi:10.1038/s41588-018-0205-x

26. Said S, Pazoki R, Karhunen V, et al. Genetic analysis of over half a million people characterises C-reactive protein loci. Nat Commun. 2022;13(1):2198. doi:10.1038/s41467-022-29650-5

27. van der Harst P, Verweij N. Identification of 64 Novel Genetic Loci Provides an Expanded View on the Genetic Architecture of Coronary Artery Disease. Circ Res. 2018;122(3):433–443. doi:10.1161/CIRCRESAHA.117.312086

28. Sakaue S, Kanai M, Tanigawa Y, et al. A cross-population atlas of genetic associations for 220 human phenotypes. Nat Genet. 2021;53(10):1415–1424. doi:10.1038/s41588-021-00931-x

29. Malik R, Chauhan G, Traylor M, et al. Multiancestry genome-wide association study of 520,000 subjects identifies 32 loci associated with stroke and stroke subtypes. Nat Genet. 2018;50(4):524–537. doi:10.1038/s41588-018-0058-3

30. Ishigaki K, Sakaue S, Terao C, et al. Multi-ancestry genome-wide association analyses identify novel genetic mechanisms in rheumatoid arthritis. Nat Genet. 2022;54(11):1640–1651. doi:10.1038/s41588-022-01213-w

31. Hemani G, Zheng J, Elsworth B, et al. The MR-Base platform supports systematic causal inference across the human phenome. Loos R, ed. eLife. 2018;7:e34408. doi:10.7554/eLife.34408

32. Burgess S, Butterworth A, Thompson SG. MR analysis with multiple genetic variants using summarized data. Genet Epidemiol. 2013;37(7):658–665. doi:10.1002/gepi.21758

33. Patel A, Ye T, Xue H, et al. MendelianRandomization v0.9.0: updates to an R package for performing MR analyses using summarized data. Wellcome Open Res. 2023;8:449. doi:10.12688/wellcomeopenres.19995.1

34. Bowden J, Davey Smith G, Haycock PC, Burgess S. Consistent Estimation in MR with Some Invalid Instruments Using a Weighted Median Estimator. Genet Epidemiol. 2016;40(4):304–314. doi:10.1002/gepi.21965

35. Bowden J, Davey Smith G, Burgess S. MR with invalid instruments: effect estimation and bias detection through Egger regression. Int J Epidemiol. 2015;44(2):512–525. doi:10.1093/ije/dyv080

36. Lefkowitz RJ. Historical review: a brief history and personal retrospective of seven-transmembrane receptors. Trends Pharmacol Sci. 2004;25(8):413–422. doi:10.1016/j.tips.2004.06.006

37. O’Dell TJ, Connor SA, Guglietta R, Nguyen PV. β-Adrenergic receptor signaling and modulation of long-term potentiation in the mammalian hippocampus. Learn Mem. 2015;22(9):461–471. doi:10.1101/lm.031088.113

38. Beaman EE, Bonde AN, Larsen SMU, et al. Blood-brain barrier permeable β-blockers linked to lower risk of Alzheimer’s disease in hypertension. Brain. 2023;146(3):1141–1151. doi:10.1093/brain/awac076

39. Gelber RP, Ross GW, Petrovitch H, Masaki KH, Launer LJ, White LR. Antihypertensive medication use and risk of cognitive impairment: The Honolulu-Asia Aging Study. Neurology. 2013;81(10):888–895. doi:10.1212/WNL.0b013e3182a351d4

40. Beitelshees AL, Zineh I. Renin–angiotensin–aldosterone system (RAAS) pharmacogenomics: implications in heart failure management. Heart Fail Rev. 2010;15(3):209–217. doi:10.1007/s10741-008-9092-z

41. Newby D, Winchester LM, Sproviero W, et al. Association between centrally active angiotensin-converting enzyme inhibitors with dementia risk. Alzheimer’s & Dementia. 2023;19(S8):e066498. doi:10.1002/alz.066498

42. Walker VM, Kehoe PG, Martin RM, Davies NM. Repurposing antihypertensive drugs for the prevention of Alzheimer’s disease: a MR study. International Journal of Epidemiology. 2020;49(4):1132–1140. doi:10.1093/ije/dyz155

43. Baird DA, Liu JZ, Zheng J, et al. Identifying drug targets for neurological and psychiatric disease via genetics and the brain transcriptome. PLOS Genetics. 2021;17(1):e1009224. doi:10.1371/journal.pgen.1009224

44. Schartum-Hansen H, Løland KH, Svingen GFT, et al. Use of Loop Diuretics is Associated with Increased Mortality in Patients with Suspected Coronary Artery Disease, but without Systolic Heart Failure or Renal Impairment: An Observational Study Using Propensity Score Matching. PLoS One. 2015;10(6):e0124611. doi:10.1371/journal.pone.0124611

