## supplemental methods for "Repurposing drugs for the prevention of vascular dementia: Evidence from drug target Mendelian randomization"

**Supplementary Methods:**

### Additional Data Sources

#### *Protein Quantitative Trait Loci Data*

All analyses were replicated using pQTL data where available. GWAS summary data from the largest scale publicly available European genome-wide association studies (GWAS) were obtained for 46 drug targets (where available) from the deCODE cohort (Ferkingstad *et al*., 2021, n=35,559)[1] and the UK Biobank PPP study (Sun *et al*., 2023, n= 54,219)[2]. Where possible, primary analyses were conducted using deCODE data, as this prevented participant overlap with all selected outcome studies, which were primarily measured in the UK Biobank. Where no deCODE data was available, UKB-PPP data was used instead. The interpretation of results from this analysis is per standard deviation (SD) increase in protein levels.

The deCODE study collected data using the using SomaScan multiplex aptamer assay (version 4). This uses protein-capture SOMAmer (Slow Offrate Modified Aptamer) reagents to bind to specific protein targets for quantification using DNA microarrays[1]. In total 4,907 aptamers representing 4,719 plasma proteins were measured with GWAS summary data available from https://www.decode.com/summarydata/. The UK Biobank Pharma Proteomics Project (UKB-PPP) study measured 2,923 plasma proteins using the Olink Explore 3072 PEA (Proximity Extension Assay). This method utilizes probes formed from antibodies and oligonucleotides generating DNA-encoded tags for measurement[2]. The pQTL data used is publicly available at https://www.synapse.org/Synapse:syn51364943/wiki/622119.

#### *Vascular Dementia Meta-Analysis*

A meta-analysis for vascular dementia was performed using the METAL toolkit[3]. This combined summary data from two recently published European GWAS. Firstly, data from MEGAVCID consortium (MEGAVCID *et al*., 2024[4]) which has N cases = 3,892, N controls = 466,606. This study combined data from 11 cohorts with VaD cases (primarily defined using International Statistical Classification of Diseases and Related Health Problems (ICD) codes) plus an additional 4 providing only controls[4]. Secondly, the FinnGen study[5] (N cases = 3,116, N controls = 433,066). The FinnGen study is a large-scale genomics initiative that has analyzed over 500,000 Finnish biobank samples and correlated genetic variation with health data to understand disease mechanisms and predispositions. The project is a collaboration between research organizations and biobanks within Finland and international industry partners. Cases were identified using the “F5_VASCDEM” endpoint which is based on the digital health record data from Finnish health registries (ICD-10: F01). An inverse-variance weighted meta-analysis was conducted to combine the studies. This gave a maximum of N= 7,008 cases and N= 899,672 controls in the meta-analysis. There was no evidence of genomic inflation in the two source GWAS (lambdas within 0.1 of 1) so no genomic control was applied during the meta-analysis.

**Additional Statistical Analysis**

All analysis was administered in R (version 4.4.0). Instrumental variable selection and harmonization was conducted using the ‘TwoSampleMR’ and ‘ieugwasr’ packages[6]. MR analysis was performed using the ‘MendelianRandomization’ R package[7, 8]. Additional sensitivity analysis was conducted using the ‘coloc’ and ‘MRlap’ R packages[9, 10, 11].

Whilst the inverse-variance weighted (IVW) MR and Wald-ratio methods (used as our primary analysis method) have the best statistical power, it does rely on stringent instrument assumptions[12]. These assume no unbalanced horizontal pleiotropy (i.e., no effect of the instruments on the outcomes through any pathway other than the drug target). Thus, the intercept in these models is fixed at zero. If there is horizontal pleiotropy (i.e. an effect of the instruments on the outcomes that does not go through the drug target, otherwise known as pre-translational pleiotropy), the effect estimate for that specific drug target will be biased. Other MR methods have more relaxed assumptions. In order to address potential violations of these assumptions additional sensitivity analysis was conducted. Weighted Median MR combines the Wald ratio estimates for each genetic variant with the median inverse-variance weight[13]. Unlike IVW MR, weighted median MR still provides a consistent estimate as long as at least 50% of the weights are from valid instruments. MR-Egger is a MR method designed to estimate the magnitude of and adjust for horizontal pleiotropy, by not constraining the intercept term to zero. Instead, the intercept estimates the average pleiotropic effect[14], and the slope provides a pleiotropy corrected causal effect estimate. Consequently, this method allows instruments to be used that are invalid under the third IVW MR assumption (exclusion restriction), and instead relies on the weaker InSIDE (Instrument Strength Independent of Direct Effect) assumption, that pleiotropic effects are independently distributed from the genetic associations with the risk factor. Both MR egger and weighted median MR were applied in addition to the IVW method (using the already selected cis-acting instruments) with the ‘MendelianRandomization’ R package[7, 8]. Both methods require 3 instrumental variables for analysis, so in many cases were unable to be performed, unless the p-value threshold was reduced from $<5x10^{-08}$. Therefore, a reduced threshold sensitivity analysis was conducted.

We re-analysed results using a reduced threshold for instrumental variable selection ($p < 5^{e-05}$, $r^2 < 0.01$ and a 1,000kb distance threshold) to enable pleiotropy robust MR methods, which require more than two SNPs as instruments. Due to low case numbers in the RA GWAS, we examined additional positive controls for anti-inflammatory targets as a sensitivity analysis. These were Crohn's disease (N cases= 12,194, N controls= 28,072)[15] and ulcerative colitis (N cases= 12,366, N controls= 33,609)[14] . To further evaluate whether LDL-C was a suitable downstream biomarker for all lipid-lowering drug targets, we examined results when using triglyceride data instead of LDL-C data from Willer et al., 2013 (N= 177,861)[16]. We also tested interleukin-6 (IL-6) instead of CRP as a downstream biomarker for anti-inflammatory targets (N= 21,758)[17]. As the blood pressure datasets are adjusted for BMI, an additional analysis was conducted using unadjusted blood pressure measurements from UK-Biobank (SBP N= 317,754 DBP N= 317,756)[18].

Overlapping samples in the exposure and outcome data in two-sample MR can result in bias[11]. In our analysis, several datasets contained data from UK Biobank; of the initial data selected as downstream biomarkers for the exposure, only the LDL-c dataset did not contain any UK Biobank participants. To assess for potential bias caused by sample overlap where applicable, 'MRlap'[11] was applied as a sensitivity analysis. MRlap leverages cross-trait LD-score regression (LDSC) to approximate the overlap between summary datasets, before performing an MR analysis. It is then able to output both the original and overlap corrected results; any difference between these two indicates possible bias introduced through sample overlap. A *cis*-acting analyses was then run for all three downstream biomarkers with UK Biobank participants (SBP, DBP & CRP) with outcomes containing UK Biobank participants (WMH, FA & MD). As this method requires 3 instrumental variables for analysis, a reduced instrument selection threshold was used ($p < 5^{e-05}$, $r^2 < 0.01$ and a 1,000kb distance threshold) in the *cis*-acting region.

***pQTL Sensitivity Analysis***

Where data were available, results were replicated using pQTLs from deCODE[1] or the UK Biobank[2]. Cis-acting instruments were selected based on a 500kb distance either side of the target gene region. Instruments were selected based on genome-wide significance and an LD clumping threshold of $r^2 < 0.001$ within a 10,000kb distance. For those pQTLs with no genome-wide significant instruments a reduced threshold of $5x10^{-5}$ was applied instead. As with the main analysis IVW/Wald ratio, weighted median and MR-Egger was applied for analysis with all five VaD outcomes.

An additional analysis including *trans*-acting instrumental variables was conducted. This can capture potential effect(s) of the drug target not occurring within the immediate gene region and provides results for drug targets where no *cis*-acting instruments were identified. It is worth noting that *trans*-acting instruments have an increased risk of horizontal pleiotropy biasing the analysis. A combined *cis* and *trans*-acting analyses were performed using pQTL summary data. Independent instruments (i.e., variants associated with target protein levels at genome-wide significance and an LD clumping threshold of $r^2 < 0.001$ within a 10,000kb distance) were selected across the whole genome. Instruments were identified for 21 drug targets for MR analysis.

#### *Statistical Colocalization Analysis*

Statistical colocalization analyses were performed to evaluate whether associations between the instrument and the drug target and the instrument and the outcomes were driven by the same causal variants using the “coloc” R package[9, 10]. Colocalization analysis tests each variant-level hypothesis with the rest (support for pairings of causal and non-causal variants) using a Bayes factor. As a Bayesian method, three informative prior probabilities are specified (that any random genetic variant in the given region is associated with trait 1, 2 or both)[9]. The method then calculates the following posterior probabilities; PP.H0: there is no association with either trait in the given region; PP.H1: there is an association with only trait 1; PP.H2: there is an association with only trait 2; PP.H3: there is an association with both traits, but they have different single causal variants; PP.H4: there is an association with both traits and they share the same single causal variant. A lack of colocalization evidence does not necessarily discount a causal effect found in MR, however it does increase the robustness of the results[19]. This was performed for both a 500kb region around each target gene as well as a 500kb region surround each instrument identified in the combined cis and trans pQTL analysis.

### Supplementary References


1. Ferkingstad E, Sulem P, Atlason BA, et al. Large-scale integration of the plasma proteome with genetics and disease. *Nat Genet*. 2021;53(12):1712-1721. doi:10.1038/s41588-021-00978-w
2. Sun BB, Chiou J, Traylor M, et al. Plasma proteomic associations with genetics and health in the UK Biobank. *Nature*. 2023;622(7982):329-338. doi:10.1038/s41586-023-06592-6
3. Willer CJ, Li Y, Abecasis GR. METAL: fast and efficient meta-analysis of genomewide association scans. *Bioinformatics*. 2010;26(17):2190-2191. doi:10.1093/bioinformatics/btq340
4. The Mega Vascular Cognitive Impairment and Dementia (MEGAVCID) consortium. A genome-wide association meta-analysis of all-cause and vascular dementia. *Alzheimer's & Dementia*. 2024;20(9):5973-5995. doi:10.1002/alz.14115
5. Kurki MI, Karjalainen J, Palta P, et al. FinnGen provides genetic insights from a well-phenotyped isolated population. *Nature*. 2023;613(7944):508-518. doi:10.1038/s41586-022-05473-8
6. Hemani G, Zheng J, Elsworth B, et al. The MR-Base platform supports systematic causal inference across the human phenome. Loos R, ed. *eLife*. 2018;7:e34408. doi:10.7554/eLife.34408
7. Burgess S, Butterworth A, Thompson SG. Mendelian randomization analysis with multiple genetic variants using summarized data. *Genet Epidemiol*. 2013;37(7):658-665. doi:10.1002/gepi.21758
8. Patel A, Ye T, Xue H, et al. MendelianRandomization v0.9.0: updates to an R package for performing Mendelian randomization analyses using summarized data. *Wellcome Open Res*. 2023;8:449. doi:10.12688/wellcomeopenres.19995.1
9. Wallace C. Eliciting priors and relaxing the single causal variant assumption in colocalisation analyses. *PLOS Genetics*. 2020;16(4):e1008720. doi:10.1371/journal.pgen.1008720
10. Giambartolomei C, Vukcevic D, Schadt EE, et al. Bayesian Test for Colocalisation between Pairs of Genetic Association Studies Using Summary Statistics. *PLOS Genetics*. 2014;10(5):e1004383. doi:10.1371/journal.pgen.1004383
11. Mounier N, Kutalik Z. Bias correction for inverse variance weighting Mendelian randomization. *Genetic Epidemiology*. 2023;47(4):314-331. doi:10.1002/gepi.22522
12. Sanderson E, Glymour MM, Holmes MV, et al. Mendelian randomization. *Nat Rev Methods Primers*. 2022;2(1):1-21. doi:10.1038/s43586-021-00092-5
13. Bowden J, Davey Smith G, Haycock PC, Burgess S. Consistent Estimation in Mendelian Randomization with Some Invalid Instruments Using a Weighted Median Estimator. *Genet Epidemiol*. 2016;40(4):304-314. doi:10.1002/gepi.21965
14. Bowden J, Davey Smith G, Burgess S. Mendelian randomization with invalid instruments: effect estimation and bias detection through Egger regression. *Int J Epidemiol*. 2015;44(2):512-525. doi:10.1093/ije/dyv080
15. de Lange KM, Moutsianas L, Lee JC, et al. Genome-wide association study implicates immune activation of multiple integrin genes in inflammatory bowel disease. *Nat Genet*. 2017;49(2):256-261. doi:10.1038/ng.3760
16. Willer CJ, Schmidt EM, Sengupta S, et al. Discovery and refinement of loci associated with lipid levels. *Nat Genet*. 2013;45(11):1274-1283. doi:10.1038/ng.2797
17. Folkersen L, Gustafsson S, Wang Q, et al. Genomic and drug target evaluation of 90 cardiovascular proteins in 30,931 individuals. *Nat Metab*. 2020;2(10):1135-1148. doi:10.1038/s42255-020-00287-2
18. UK Biobank. Neale lab. Accessed March 21, 2025. http://www.nealelab.is/uk-biobank

**19.** Zuber V, Grinberg NF, Gill D, et al. Combining evidence from Mendelian randomizationMR and colocalization: Review and comparison of approaches. *Am J Hum Genet*. 2022;109(5):767-782. doi:10.1016/j.ajhg.2022.04.001
