## supplemental figures for "Repurposing drugs for the prevention of vascular dementia: Evidence from drug target Mendelian randomization"

**Supplementary Figure 1:** Additional Plots for Lipid Lowering Drug Target MR using LDL Downstream Biomarker Data, for which there was no reductive effect on the positive control

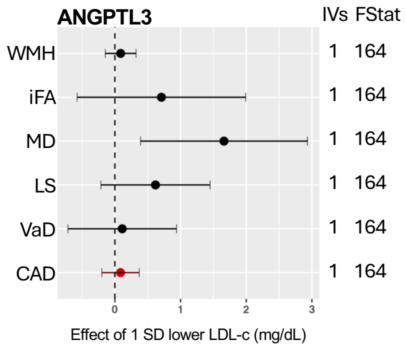


*Additional plots for the lipid-lowering drug target Mendelian randomization using LDL-c (a downstream biomarker) as the exposure with five vascular dementia outcomes; WMH (white matter hyperintensities), iFA (inversed fractional anisotropy), MD (mean diffusivity), LS (lacunar stroke), VaD (vascular dementia diagnosis). CAD (coronary artery disease) was used as a positive control and is indicated in red, the additional targets presented here did not show the expected lowering effect on the positive control (p>0.1). The number of instrumental variables (IVs) used is also given along with the F-statistic (Fstat). LDL-c (low-density lipoprotein cholesterol), SD (standard deviation).*

**Supplementary Figure 2:** Additional Plots for Antihypertensive Drug Target MR using DBP Downstream Biomarker Data , for which there was no reductive effect found on the positive control

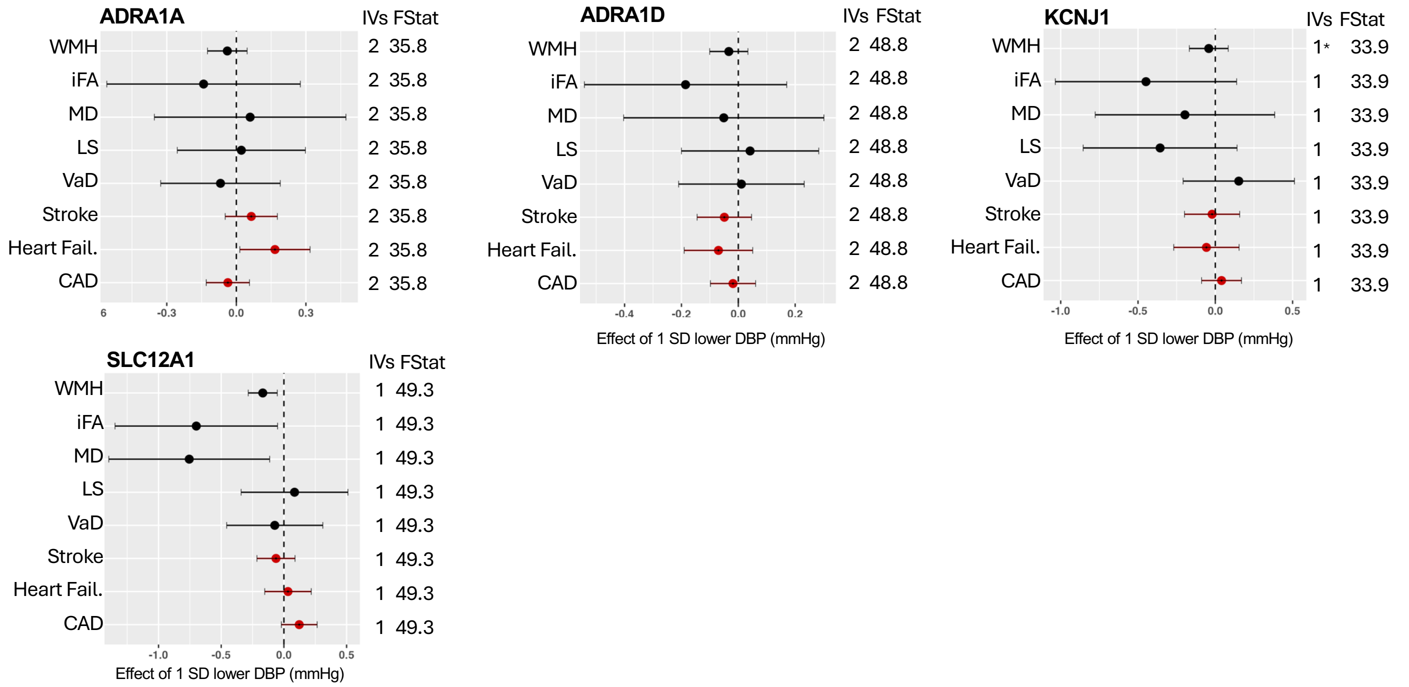


*Additional plots for the antihypertensive drug target Mendelian randomization using DBP (a downstream biomarker) as the exposure with five vascular dementia outcomes; WMH (white matter hyperintensities), iFA (inversed fractional anisotropy), MD (mean diffusivity), LS (lacunar stroke), VaD (vascular dementia diagnosis). CAD (coronary artery disease); heart disease (Heart Fail.) and stroke were used as a positive control and are indicated in red; the additional targets presented here did not show the expected lowering effect on the positive control (p>0.1). The number of instrumental variables (IVs) used is also given along with the F-statistic (Fstat). If the WMH meta-analysis dataset could not be used, the UK-Biobank only WMH dataset is presented instead (indicated by a * next to the IVs). DBP (diastolic blood pressure), SD (standard deviation).*

**Supplementary Figure 3:** Additional Plots for Antihypertensive Drug Target MR using SBP Downstream Biomarker Data

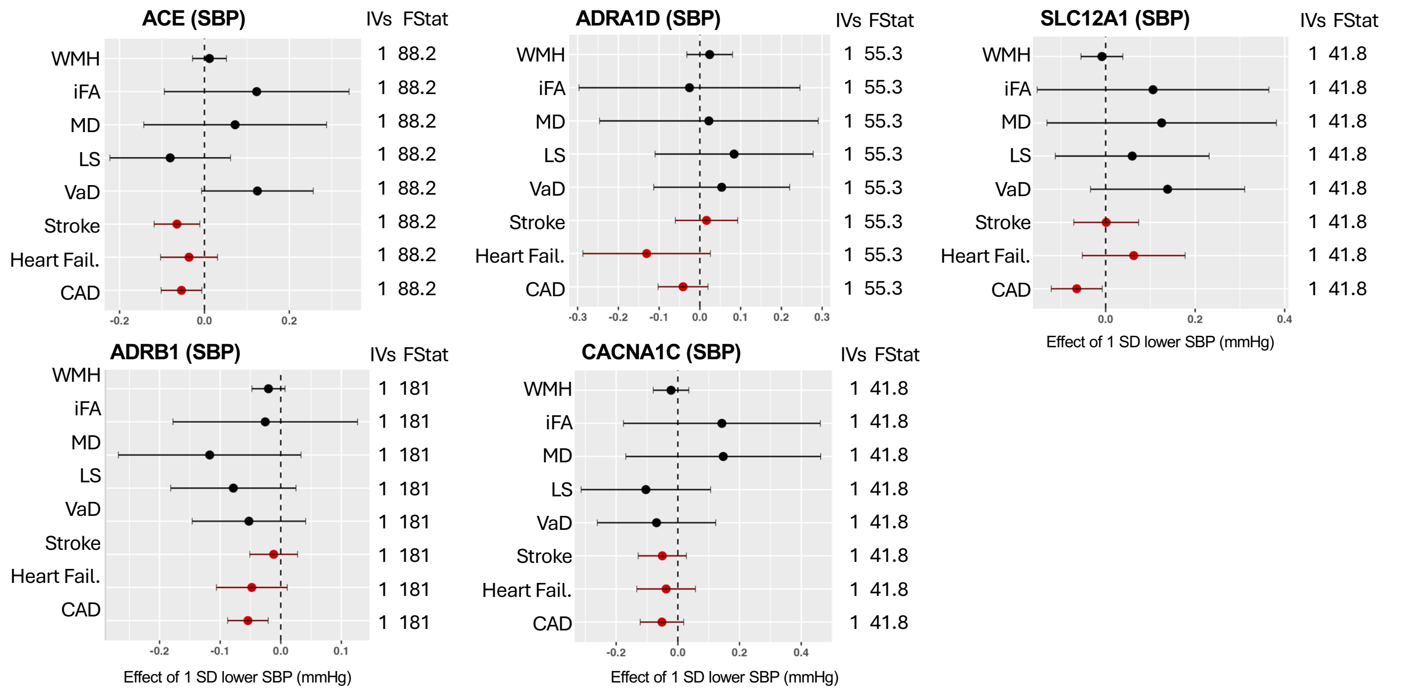


*For the antihypertensive drug targets which had instruments identified for both DBP and SBP downstream biomarkers the plots for the Mendelian randomization using SBP as the exposure with five vascular dementia outcomes; WMH (white matter hyperintensities), iFA (inversed fractional anisotropy), MD (mean diffusivity), LS (lacunar stroke), VaD (vascular dementia diagnosis) are given here. CAD (coronary artery disease); heart disease (Heart Fail.) and stroke were used as a positive control and are indicated in red. The number of instrumental variables (IVs) used is also given along with the F-statistic (Fstat). SBP (systolic blood pressure), SD (standard deviation).*

**<u>Supplementary Figure 4a:</u>** <u>Additional Plots for Anti-inflammatory Drug Target MR using CRP Downstream Biomarker Data, for which there was no reductive effect found on the positive control</u>

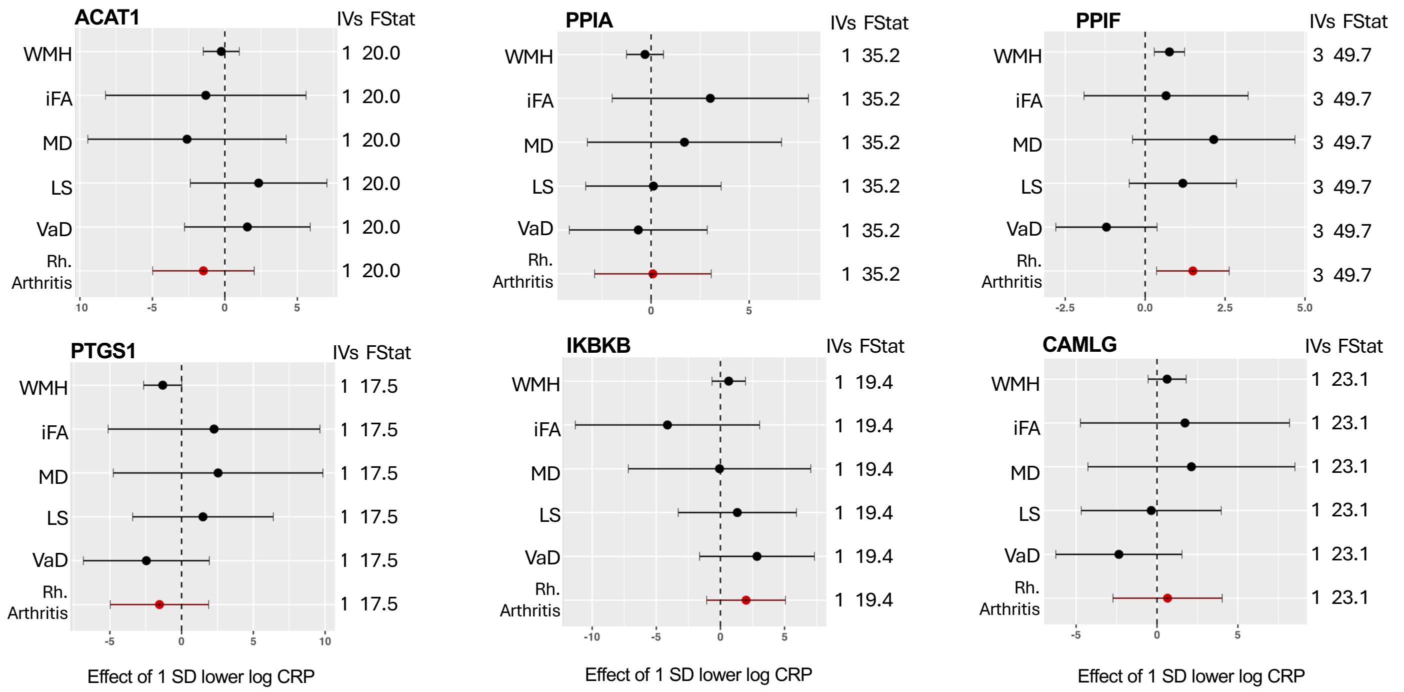


*Additional plots for the antihypertensive drug target Mendelian randomization using CRP (a downstream biomarker) as the exposure with five vascular dementia outcomes; WMH (white matter hyperintensities), iFA (inversed fractional anisotropy), MD (mean diffusivity), LS (lacunar stroke), VaD (vascular dementia diagnosis). Rheumatoid arthritis (Rh. Arthritis) was used as a positive control and is indicated in red; the additional targets presented here did not show the expected lowering effect on the positive control (p>0.1). The number of instrumental variables (IVs) used is also given along with the F-statistic (Fstat). CRP (c-reactive protein), SD (standard deviation).*

**Supplementary Figure 4b:** Additional Plots for Anti-inflammatory Drug Target MR using CRP Downstream Biomarker Data, for which there was no reductive effect found on the positive control

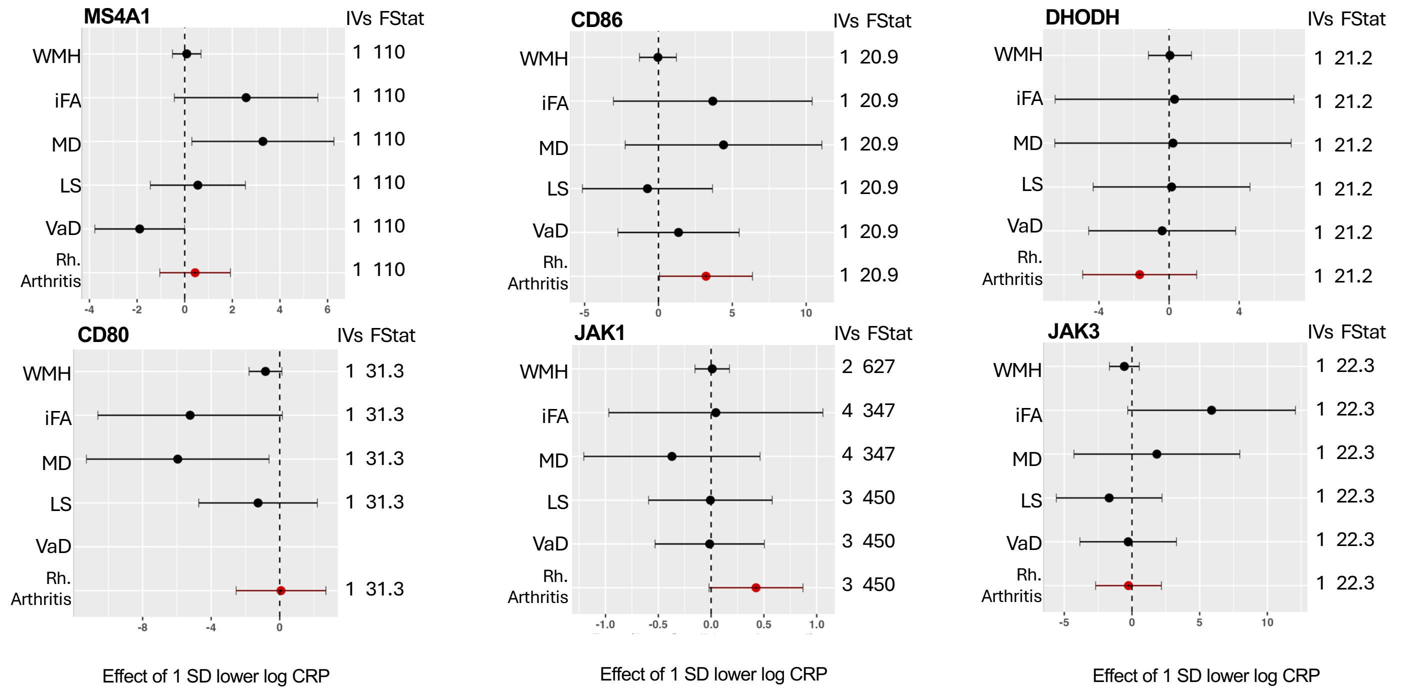


*Additional plots for the antihypertensive drug target Mendelian randomization using CRP (a downstream biomarker) as the exposure with five vascular dementia outcomes; WMH (white matter hyperintensities), iFA (inversed fractional anisotropy), MD (mean diffusivity), LS (lacunar stroke), VaD (vascular dementia diagnosis). Rheumatoid arthritis (Rh. Arthritis) was used as a positive control and is indicated in red; the additional targets presented here did not show the expected lowering effect on the positive control (p>0.1). The number of instrumental variables (IVs) used is also given along with the F-statistic (Fstat). CRP (c-reactive protein), SD (standard deviation).*

**Supplementary Figure 4c:** Additional Plots for Anti-inflammatory Drug Target MR using CRP Downstream Biomarker Data, for which there was no reductive effect found on the positive control

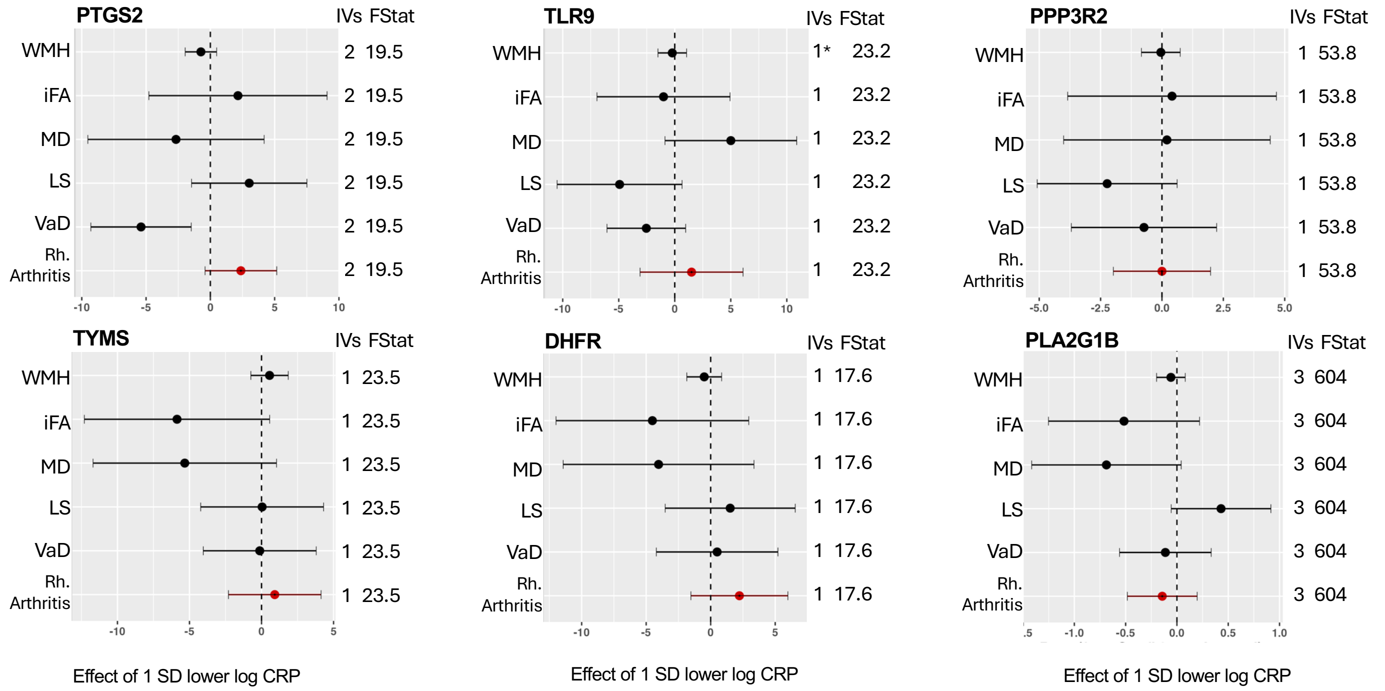


*Additional plots for the antihypertensive drug target Mendelian randomization using CRP (a downstream biomarker) as the exposure with five vascular dementia outcomes; WMH (white matter hyperintensities), iFA (inversed fractional anisotropy), MD (mean diffusivity), LS (lacunar stroke), VaD (vascular dementia diagnosis). Rheumatoid arthritis (Rh. Arthritis) was used as a positive control and is indicated in red; the additional targets presented here did not show the expected lowering effect on the positive control (p>0.1). The number of instrumental variables (IVs) used is also given along with the F-statistic (Fstat). CRP (c-reactive protein), SD (standard deviation).*

**Supplementary Figure 5:** Plots for Lipid-Lowering Drug Target MR using cis-pQTL Data

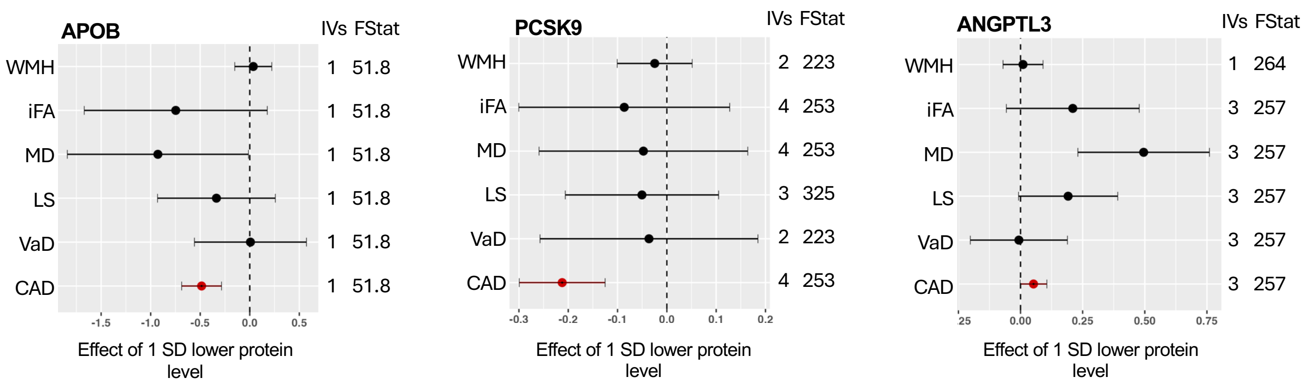


*Plots for the lipid-lowering drug target Mendelian randomization using cis-acting pQTLs as the exposure with five vascular dementia outcomes; WMH (white matter hyperintensities), iFA (inversed fractional anisotropy), MD (mean diffusivity), LS (lacunar stroke), VaD (vascular dementia diagnosis). CAD (coronary artery disease) was used as a positive control and is indicated in red. The number of instrumental variables (IVs) used is also given along with the F-statistic (Fstat). SD (standard deviation).*

**Supplementary Figure 6:** Plots for Anti-hypertensive Drug Target MR using cis-pQTL Data

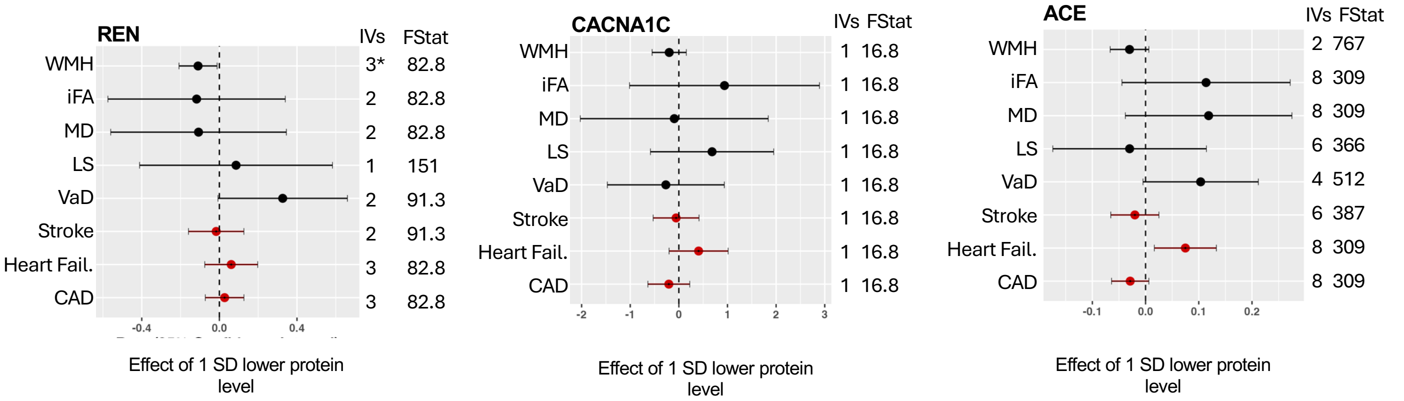


*Plots for the antihypertensive drug target Mendelian randomization using cis-acting pQTLs as the exposure with five vascular dementia outcomes; WMH (white matter hyperintensities), iFA (inversed fractional anisotropy), MD (mean diffusivity), LS (lacunar stroke), VaD (vascular dementia diagnosis). CAD (coronary artery disease); heart disease (Heart Fail.) and stroke were used as a positive control and are indicated in red. The number of instrumental variables (IVs) used is also given along with the F-statistic (Fstat). If the WMH meta-analysis dataset could not be used, the UK-Biobank only WMH dataset is presented instead (indicated by a * next to the IVs). SD (standard deviation), pQTL (protein quantitative trait loci).*

**Supplementary Figure 7a:** Plots for Anti-inflammatory Drug Target MR using cis-pQTL Data

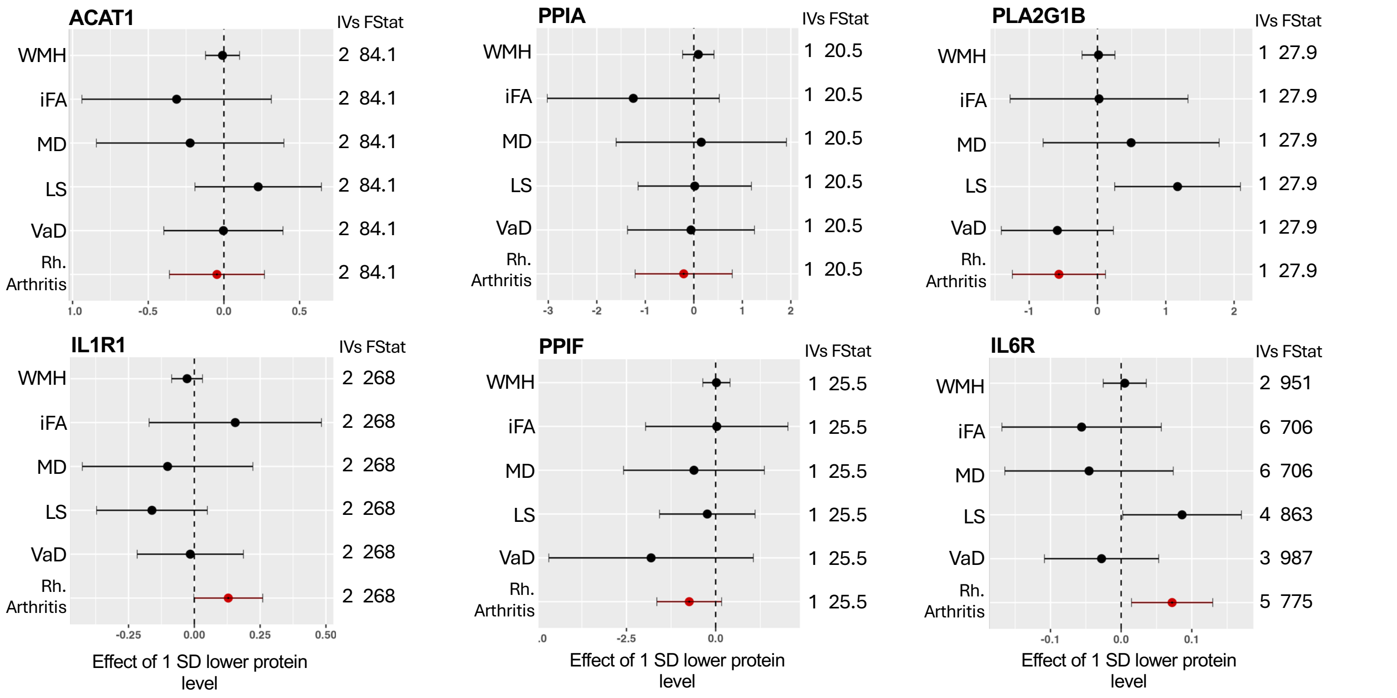


*Plots for the antihypertensive drug target Mendelian randomization using cis-acting pQTLs as the exposure with five vascular dementia outcomes; WMH (white matter hyperintensities), iFA (inversed fractional anisotropy), MD (mean diffusivity), LS (lacunar stroke), VaD (vascular dementia diagnosis). Rheumatoid arthritis (Rh. Arthritis) was used as a positive control and is indicated in red The number of instrumental variables (IVs) used is also given along with the F-statistic (Fstat). SD (standard deviation), pQTL (protein quantitative trait loci).*

**Supplementary Figure 7b:** Plots for Anti-inflammatory Drug Target MR using cis-pQTL Data

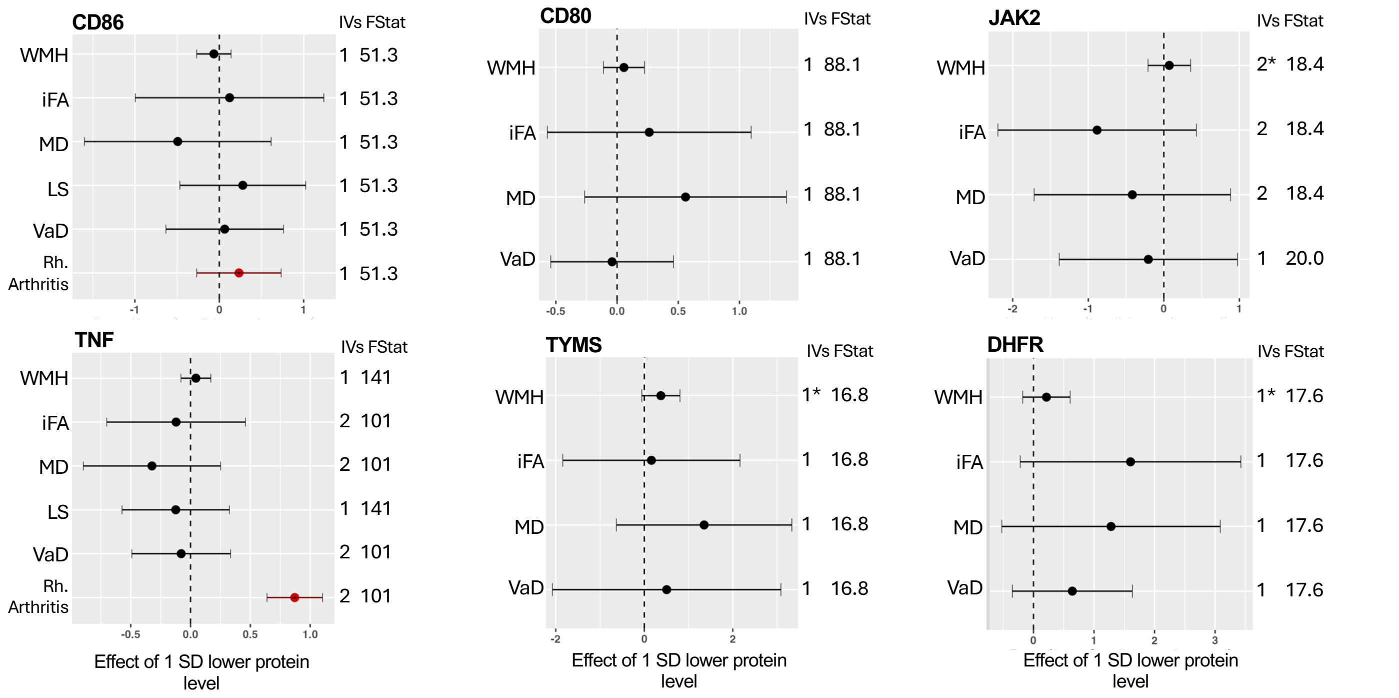


*Plots for the anti-inflammatory drug target Mendelian randomization using cis-acting pQTLs as the exposure with five vascular dementia outcomes; WMH (white matter hyperintensities), iFA (inversed fractional anisotropy), MD (mean diffusivity), LS (lacunar stroke), VaD (vascular dementia diagnosis). Rheumatoid arthritis (Rh. Arthritis) was used as a positive control and is indicated in red The number of instrumental variables (IVs) used is also given along with the F-statistic (Fstat). If the WMH meta-analysis dataset could not be used, the UK-Biobank only WMH dataset is presented instead (indicated by a * next to the IVs). SD (standard deviation), pQTL (protein quantitative trait loci).*

**Supplementary Figure 8:** <u>Plots for Lipid-Lowering Drug Target MR using *cis* and *trans* pQTL Instruments for Analysis</u>

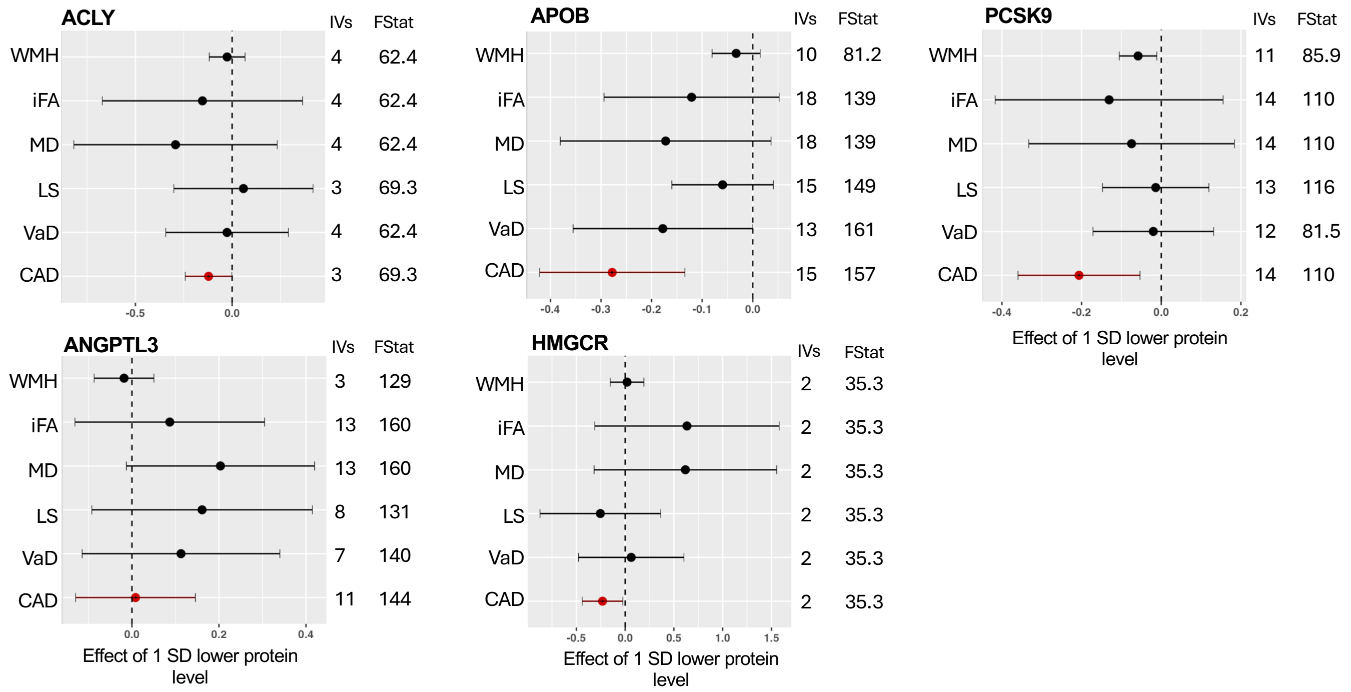


*Plots for the lipid-lowering drug target Mendelian randomization using both cis-acting and trans-acting pQTLs as the exposure with five vascular dementia outcomes; WMH (white matter hyperintensities), iFA (inversed fractional anisotropy), MD (mean diffusivity), LS (lacunar stroke), VaD (vascular dementia diagnosis). CAD (coronary artery disease) was used as a positive control and is indicated in red. The number of instrumental variables (IVs) used is also given along with the F-statistic (Fstat). SD (standard deviation), pQTL (protein quantitative trait loci).*

**Supplementary Figure 9:** Plots for Anti-hypertensive Drug Target MR using *cis* and *trans* pQTL Instruments for Analysis

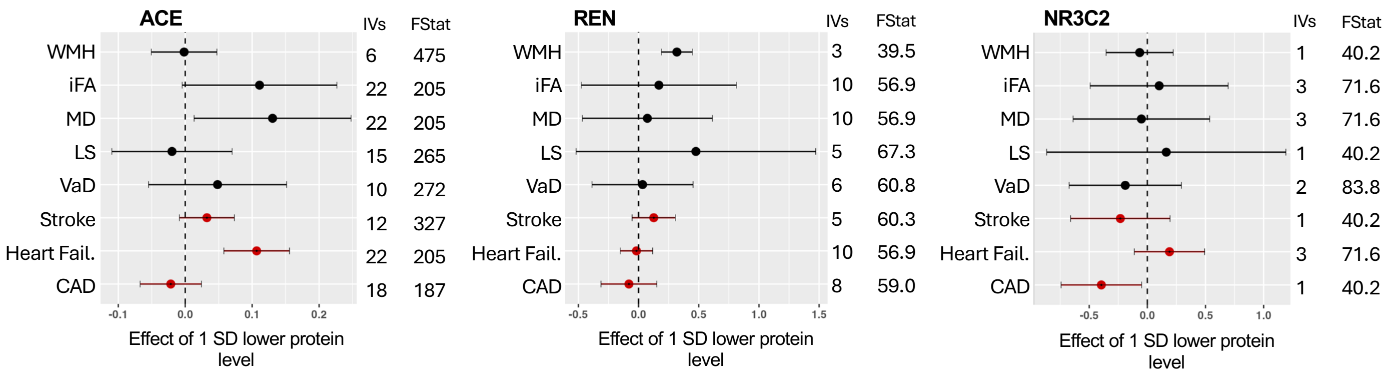


*Plots for the antihypertensive drug target Mendelian randomization using cis-acting and trans-acting pQTLs as the exposure with five vascular dementia outcomes; WMH (white matter hyperintensities), iFA (inversed fractional anisotropy), MD (mean diffusivity), LS (lacunar stroke), VaD (vascular dementia diagnosis). CAD (coronary artery disease); heart disease (Heart Fail.) and stroke were used as a positive control and are indicated in red. The number of instrumental variables (IVs) used is also given along with the F-statistic (Fstat). SD (standard deviation), pQTL (protein quantitative trait loci).*

**Supplementary Figure 10a:** Plots for Anti-inflammatory Drug Target MR using *cis* and *trans* pQTL Instruments for Analysis

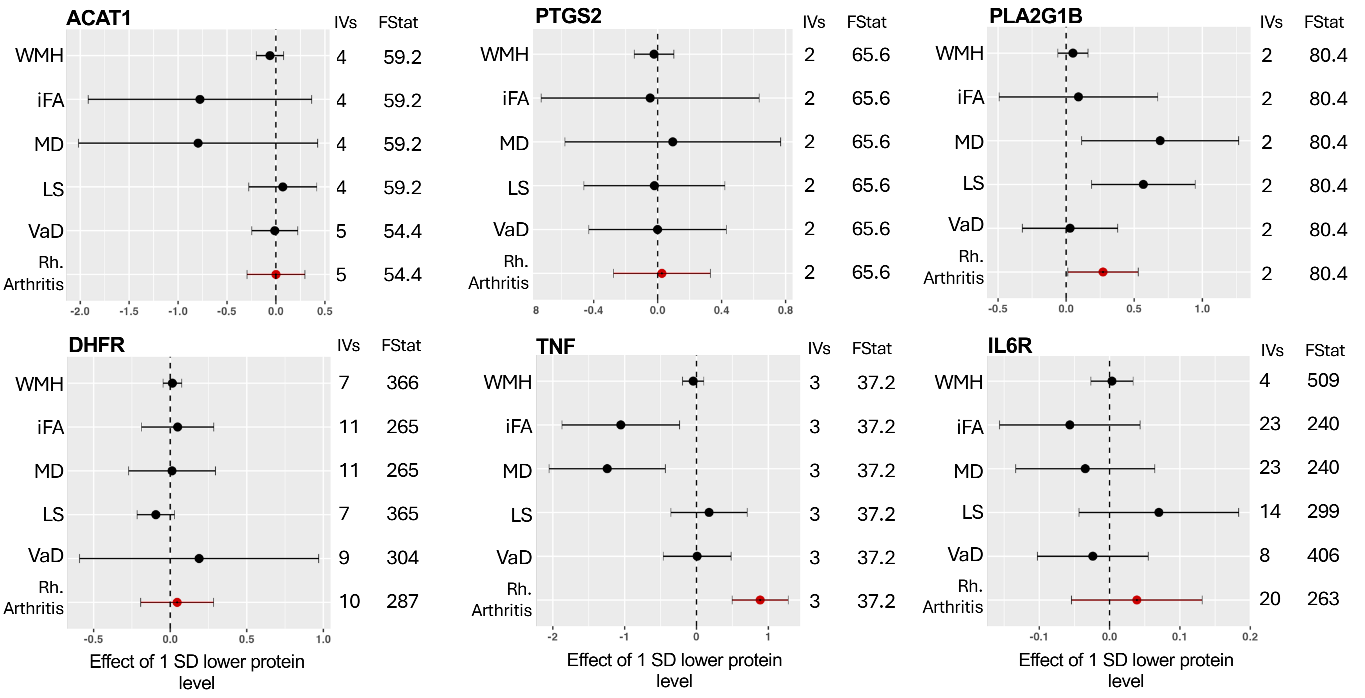


*Plots for the anti-inflammatory drug target Mendelian randomization using cis-acting and trans-acting pQTLs as the exposure with five vascular dementia outcomes; WMH (white matter hyperintensities), iFA (inversed fractional anisotropy), MD (mean diffusivity), LS (lacunar stroke), VaD (vascular dementia diagnosis). Rheumatoid arthritis (Rh. Arthritis) was used as a positive control and is indicated in red The number of instrumental variables (IVs) used is also given along with the F-statistic (Fstat). SD (standard deviation), pQTL (protein quantitative trait loci).*

**Supplementary Figure 10b:** Plots for Anti-inflammatory Drug Target MR using *cis* and *trans* pQTL Instruments for Analysis

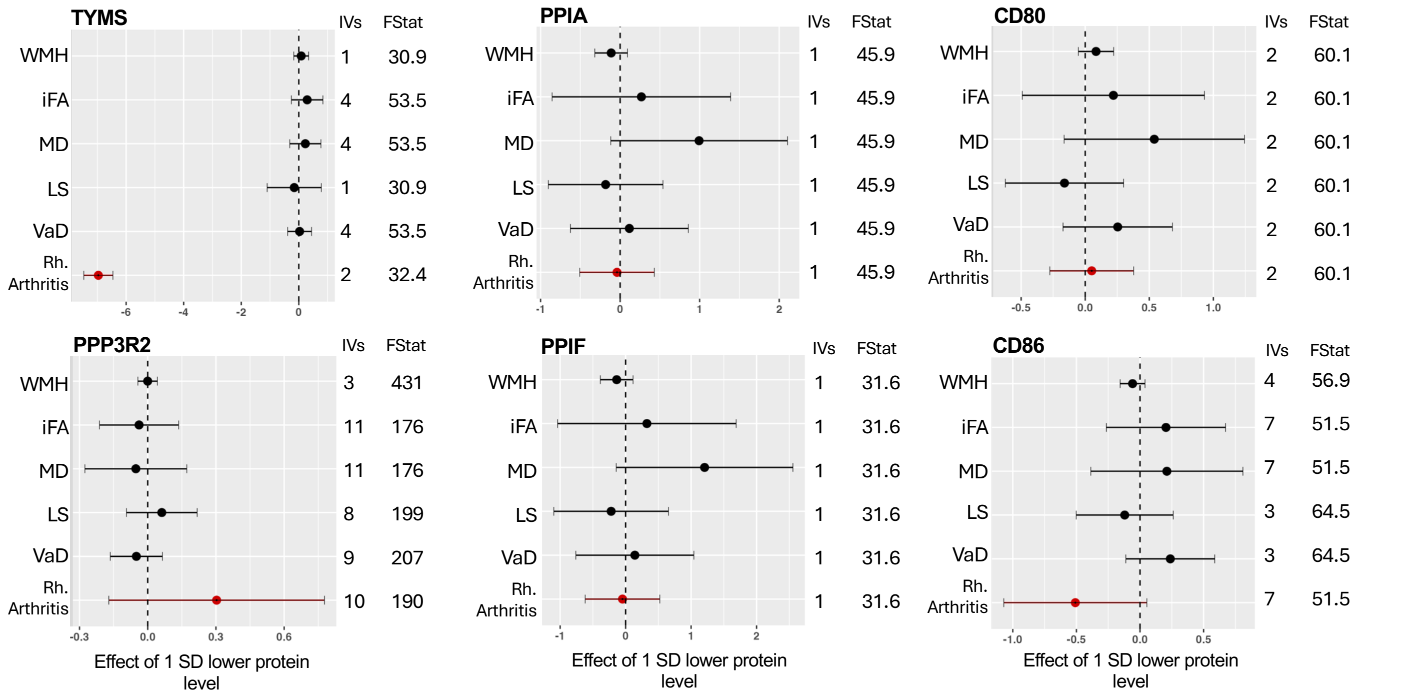


*Plots for the anti-inflammatory drug target Mendelian randomization using cis-acting and trans-acting pQTLs as the exposure with five vascular dementia outcomes; WMH (white matter hyperintensities), iFA (inversed fractional anisotropy), MD (mean diffusivity), LS (lacunar stroke), VaD (vascular dementia diagnosis). Rheumatoid arthritis (Rh. Arthritis) was used as a positive control and is indicated in red The number of instrumental variables (IVs) used is also given along with the F-statistic (Fstat). SD (standard deviation), pQTL (protein quantitative trait loci).*

**Supplementary Figure 10c:** Plots for Anti-inflammatory Drug Target MR using *cis* and *trans* pQTL Instruments for Analysis

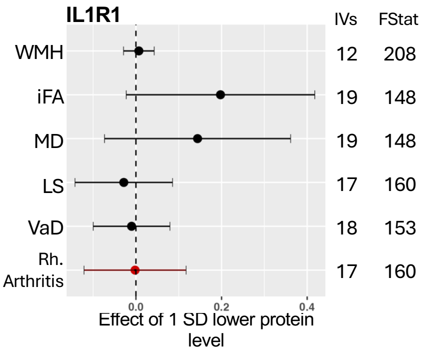


*Plots for the anti-inflammatory drug target Mendelian randomization using cis-acting and trans-acting pQTLs as the exposure with five vascular dementia outcomes; WMH (white matter hyperintensities), iFA (inversed fractional anisotropy), MD (mean diffusivity), LS (lacunar stroke), VaD (vascular dementia diagnosis). Rheumatoid arthritis (Rh. Arthritis) was used as a positive control and is indicated in red The number of instrumental variables (IVs) used is also given along with the F-statistic (Fstat). SD (standard deviation), pQTL (protein quantitative trait loci).*
